# Data Simulation to Optimize the GWAS Framework in Diverse Populations

**DOI:** 10.1101/2023.10.26.23297606

**Authors:** Jacquiline Wangui Mugo, Emile Rugamika Chimusa, Nicola Mulder

## Abstract

Whole-genome or genome-wide association studies have become a fundamental part of modern genetic studies and methods for dissecting the genetic architecture of common traits based on common polymorphisms in random populations. It is hoped that there will be many potential uses of these identified variants, including a better understanding of the pathogenesis of traits, the discovery of biomarkers and protein targets, and the clinical prediction of drug treatments for global health. Questions have been raised on whether associations that are largely discovered in populations of European descent are replicable in diverse populations, can inform medical decision-making globally, and how efficiently current GWAS tools perform in populations of high genetic diversity, multi-wave genetic admixture, and low linkage disequilibrium (LD), such as African populations. In this study, we employ genomic data simulation to mimic structured African, European, and multi-way admixed populations to evaluate the replicability of association signals from current state-of-the-art GWAS tools in these populations. We then leverage the results to discuss an optimized framework for the analysis of GWAS data in diverse populations and outline the implications, challenges, and opportunities these studies present for populations of non-European descent.

## Introduction

The frequent occurrence of population differences in phenotype outcomes and drug responses has important consequences for the biomedical sciences and industries. This has been shown to be a result of variation in host genomes and differing environments (Evangelou and Ioannidis, 2013; Goetz et al., 2014). For over a decade, GWAS have been used successfully for detecting variants in LD within causal genes (Visscher et al., 2017). These approaches have become a fundamental part of modern genetic studies, and methods for dissecting the genetic architecture of common traits based on common polymorphisms in different populations have been developed (Purcell et al., 2007; Yang et al., 2011; Seldin et al., 2011; Loh et al., 2015). As a result, our knowledge of the genetic architecture of complex diseases, such as heritability estimation, the genetic correlation between diseases, the number of loci, and their effect sizes, has been enhanced (Zaitlen et al., 2014; Brody et al., 2017; Chimusa et al., 2019; Duncan et al., 2019).

Thus far, many new genetic associations with diseases have been identified (Buniello et al., 2019). However, common current approaches to identifying the associations have been mostly designed to capture genomes with a long range of LD and haplotypes, such as those found in populations of European descent that have mostly undergone a population bottleneck (Martin et al., 2017). Consequently, GWAS today continues to be dominated by studies conducted on European cohorts (Sirugo et al., 2019). A concerning observation is that large numbers of modern drugs approved by the Food and Drug Administration (FDA) and similar organizations have been developed with relevance to Caucasian ancestry populations, yet research continues to reveal that subtle differences in the genetic make-up of other populations, such as Asian, South American, and African populations, can affect treatment (Petrovski and Goldstein, 2016; Sirugo et al., 2019). This is evidenced by the hundreds of thousands of deaths occurring annually due to adverse drug reactions resulting from differing factors, including disease determinants, environmental exposure, the human microbiome profile, and genetic factors (Hassan et al., 2021). The use of genetic information to inform medical decision-making therefore raises questions as to whether such use could be equitable. It is, therefore, crucial to extend GWAS to diverse global population cohorts as well as assess how well current approaches capture associations in these populations.

Given differences in allelic architecture, the differing pattern of LD, and the confounding of environmental factors across populations, the richer mixtures of diverse population genetic variants and differing environments are likely to contribute to a wider phenotypic variability (Campbell and Tishkoff, 2008; Sirugo et al., 2019).

Significant effort has been invested in designing GWAS mapping tools since the first GWAS was conducted. Researchers have explored various models and technologies, which have resulted in GWAS tools with better power, improved efficiency, and a significantly lower computational cost. In particular, linear mixed models (LMMs) have become an attractive approach in GWAS due to their effectiveness in capturing possible population structures in the data. Other common approaches for population structure control include the use of genomic controls (GC), the inclusion of principal components (PCs) as covariates, and structured associations (Hellwege et al., 2017). However, LMMs implement a genetic relatedness matrix (GRM) in the calculation of phenotypic variance, which allows these approaches to capture a wide range of structures, from cryptic relatedness to population stratification (Korte et al., 2012; Korte and Farlow, 2013). As inclusion of diverse populations into GWAS is underway, it is therefore not surprising that LMM has become the go-to method of choice for many researchers in GWAS analysis of these populations (Chimusa et al., 2014; Chung and Zou, 2014; Conomos et al., 2016; Swenson et al., 2018). This is often supplemented with a control for global ancestry using PC axes as covariates and GC applied to the summary statistics. A substantial number of GWAS in admixed African Americans, Latinos, and African populations today have been conducted using LMM approaches (Hoffman, 2013; Chimusa et al., 2014; Burkart et al., 2018; Daya et al., 2019), however, other GWAS methods, including logistic regression (Chen et al., 2015) has also been employed in these populations.

Though LMM-based tools have become a standard in GWAS analysis, it has been noted that they do not fully control for sub-variant structure between affected and unaffected samples, especially if there is an environmental component to phenotypic associations with ancestry at local variants or locus-specific ancestry due to admixture (Winkler et al., 2010; Seldin et al., 2011; Brody et al., 2017; Visscher et al., 2017). Non-genetic factors, such as environmental exposures, may be correlated with genetic ancestry due to the shared local environment (familial or community effects) or due to the relationship between ancestry and socio-cultural factors such as ethnicity and religious background (Mcgrath et al., 2013). Effective methods are thus needed to both leverage and control the effect of local-specific ancestry tracts in variant-level GWAS, which may further improve power and reduce false positives in mixed or multi-ancestry samples (Mcgrath et al., 2013; Marigorta et al., 2018; Awany et al., 2019).

This study leverages realistic and robust simulations that mimick European, African, and admixed populations to investigate how well current commonly used state-of-the-art GWAS analysis tools capture disease signals of similar strength in the different populations, given that most GWAS tools are benchmarked using the European population. We identify the challenges and provide an overview of the prospects for individualized prediction of disease risk and its foreseeable impact on clinical practice in people of non-European descent.

## Methods

### Leveraging Data Simulation Framework for GWAS Analysis in Diverse Populations

Simulation of homogeneous and admixed case-control populations with well-known structures that mimic real populations may help to better understand their genetic variations and evaluate different existing GWAS tools for complex disease association analysis. The genetic structure of populations as well as other controllable factors, including allele frequency and LD patterns of genetic markers, are important in the simulation of genotype data for GWAS. It is important to note that the power of a statistical test to detect a risk locus relies heavily on the allelic spectrum (numbers and frequencies of alleles) and the LD structure around the locus. Therefore, simulated data should possess both local and long-range LD (LRLD) patterns and maintain allele frequencies like real data (Ripke et al., 2015). The resampling approach starts with real data and avoids the use of an evolutionary process. It has been shown that this method, compared to other approaches, has the advantage of retaining real data properties such as allele frequency and LD in the initial pool of data (Li and Stephens, 2003; Ripke et al., 2015).

To facilitate the assessment of common GWAS tools, we simulated homogeneous and heterogeneous datasets based on haplotypes from the 1000 Genomes project spanning the genome and realistic enough to mimic African, European, and admixed populations to challenge the statistical methods for association testing in real-world conditions. We used a resampling model with recombination breakpoints while mimicking mutation rates as implemented in FractalSIM (Mugo et al., 2017).

The African and European populations were simulated under a homogeneous simulation model. We merged five European and two West African populations to form the reference population for the simulation. The merged populations, the corresponding sample sizes, and the abbreviations for the populations used are listed in **Table 1**.

**Table 1:** The table lists the different populations, their abbreviations, and the corresponding sample sizes for the populations merged to obtain the reference population for the simulation of European and African populations.

| **Simulation** | **Reference Population** | **Sample Size** |
| --- | --- | --- |
| European | British (GBR) | 85 |
|  | Iberian Spanish (IBS) | 107 |
|  | Finnish (FIN) | 99 |
|  | Toscani in Italy (TSI) | 104 |
|  | Utah residents with Northern and Western European ancestry (CEU) | 94 |
| **Merged European reference** |  | **489** |
| African | Gambian Mandinka (GWD) | 113 |
|  | Yoruba (YRI) | 108 |
| **Merged African reference** |  | **221** |

We selected 9,139,969 similar biallelic SNPs in the European and African populations. Two sets of case-control datasets with an equal number of cases and controls (500 cases, 500 controls and 2500 cases, 2500 controls) were simulated for each merged population. These sample sizes were chosen to allow a realistic evaluation of GWAS power for the different tools in European versus African populations, as most GWAS in non-European populations still suffer from small sample sizes.

For each of the simulations, in each of the sample sizes, for both the European and African populations, we selected a total of 6 SNPs to be simulated with causal effects. The SNPs were selected to be spread across the genome, and as such, we chose risk SNPs on chromosomes 2, 6, 11, 15, and 20. **Figure 1** illustrates our choice of the risk SNPs on the different chromosomes. On chromosome 2, we chose 2 SNPs, *rs*113456069 and *rs*112486568, that were selected such that they were in complete LD (*r*^2^ = 1) in the European dataset. SNP *rs*113456069 was then simulated as causal in the European population, while *rs*112486568 was simulated as causal in the African population. Both SNPs were simulated with the same signal strength in both populations. A similar process was applied in choosing the causal SNPs on chromosome 20. SNPs *rs*6115358 and *rs*7343318 were in complete LD in the European reference dataset, but only *rs*6115358 was simulated as causal in the European population and *rs*57343318 simulated as causal in the African populations. The objective of this design in choosing the causal SNPs on chromosomes 2 and 20 was to enable investigation of the replicability of GWAS results observed in the European population in an African population GWAS study using different tools, given that the risk variants in the African GWAS and in the European GWAS, are in LD in the European GWAS. The causal SNPs on chromosome 11 were chosen such that they were in complete LD in both populations, but in the European population they were simulated to have a strong signal, while in the African population they were simulated to have a weak signal. This design was to enable the investigation of different tools for capturing disease signals in the GWAS of the African population as the sample size increased when the signal strength was weak. On chromosomes 6 and 15, both SNPs were simulated with the same signal strength in both populations.

**Figure 1:**
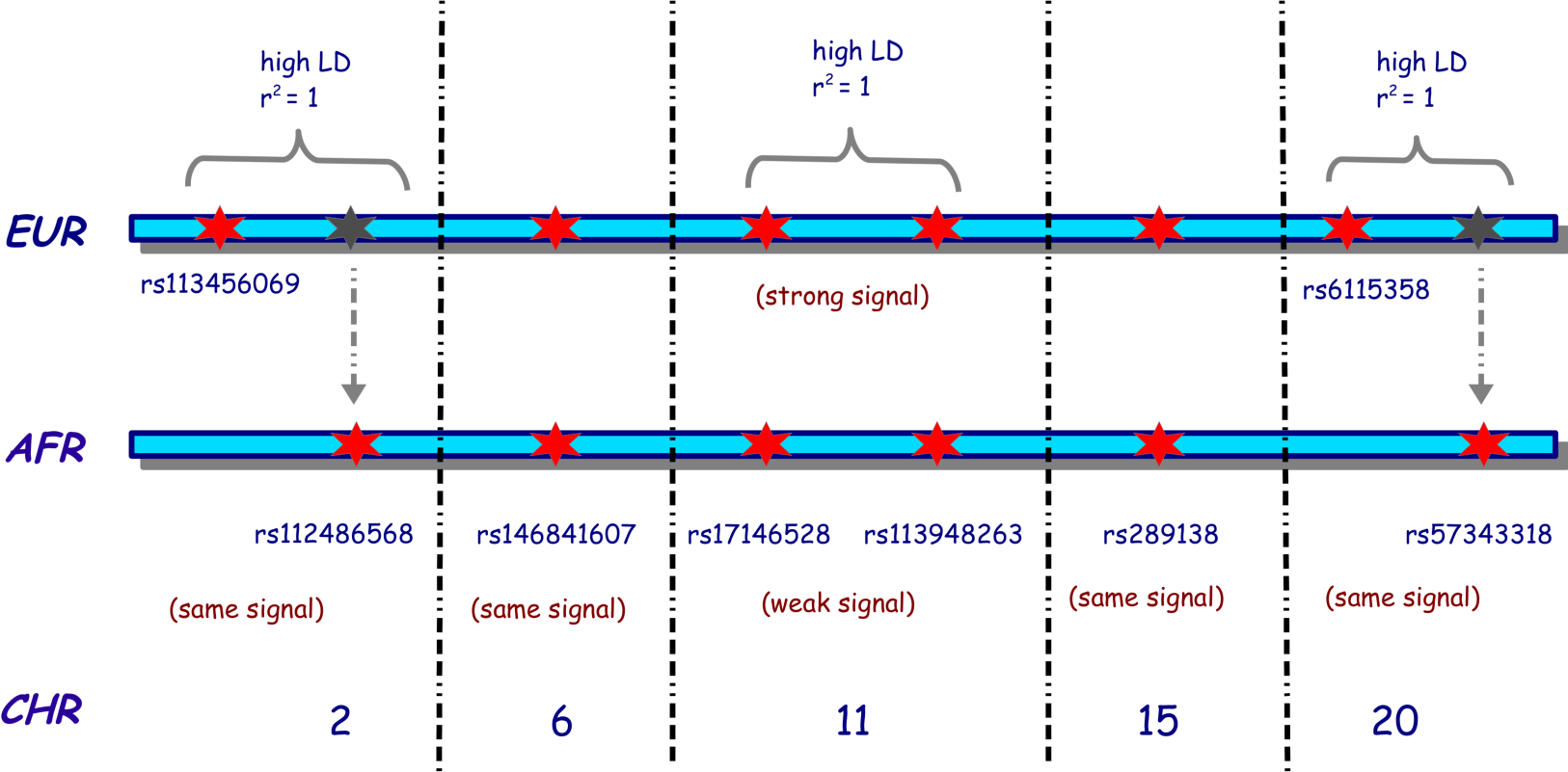
The figure illustrates the choice of the risk SNPs selected for the simulation of homogeneous European and African populations. EUR indicates European, AFR indicates African, and CHR indicates the chromosome. A red star indicates the SNP was simulated as a risk SNP in that population. In contrast, a grey star indicates a SNP is present in the European population, in high LD with the risk SNP simulated on that chromosome, but simulated as a risk SNP in the African Population. The black dotted vertical lines indicate the presence of other chromosomes between the two chromosomes. The risk SNPs on chromosomes 2, 6, 15, and 20 were simulated with the same risk strength in both populations, while on chromosome 11, the risk SNPs of Europeans were simulated with a strong risk signal, while those of Africans were simulated with a weak risk signal.

We specified the same homozygosity and heterozygosity relative risks for the 8 risk SNPs for both the 500 cases, 500 controls and 2500 cases, 2500 controls simulations. The list of these SNPs and the corresponding relative risks in the European and African population simulations are listed in **Table 2**. The cases and controls were then simulated using a multiple logistic regression model implemented in FractalSIM.

**Table 2:** The table lists the simulated risk SNPs and their corresponding homozygosity (HOM) and heterozygosity (HET) risks specified during simulation; - indicates the SNP was not simulated in that population.

| Chr | rsID | Position | European Population |  | African Population |  |
| --- | --- | --- | --- | --- | --- | --- |
|  |  |  | HOM | HET | HOM | HET |
| 2 | rs113456069 | 113842451 | 2.5004 | 0.124 | - | - |
| 2 | rs112486568 | 113842455 | - | - | 2.5004 | 0.124 |
| 6 | rs146841607 | 29942575 | 2.5004 | 0.051 | 2.5004 | 0.051 |
| 11 | rs17146528 | 64732006 | 1.5004 | 0.304 | 1.5004 | 0.704 |
| 11 | rs113948263 | 64733224 | 1.5004 | 0.304 | 1.5004 | 0.704 |
| 15 | rs289138 | 62599775 | 1.5004 | 0.104 | 1.5004 | 0.104 |
| 20 | rs6115358 | 25856361 | 1.5004 | 0.304 | - | - |
| 20 | rs7343318 | 25856699 | - | - | 0.5004 | 0.304 |

The heterogeneous datasets were generated under a single-point admixture scenario, where the admixture process occurs at a single point in history, such that the current generation is the offspring of the admixed population that has interbred over the years. Considering a random mating model where interbreeding has occurred for 10 generations, the admixture simulation first mimicked the isolated growth of each population, where a disease model (causal or null) was simulated in the isolated homogeneous simulation for each of the parental populations, similar to the case-control homogeneous simulation of the European and African populations detailed above. At generation 0, the isolated populations were allowed to interbreed. We simulated both 3-way and 5-way admixture scenarios. **Table 3** lists the reference parental populations used in the 2 scenarios, their corresponding initial sample sizes, and the proportion of ancestry contribution of each of the populations.

**Table 3:**
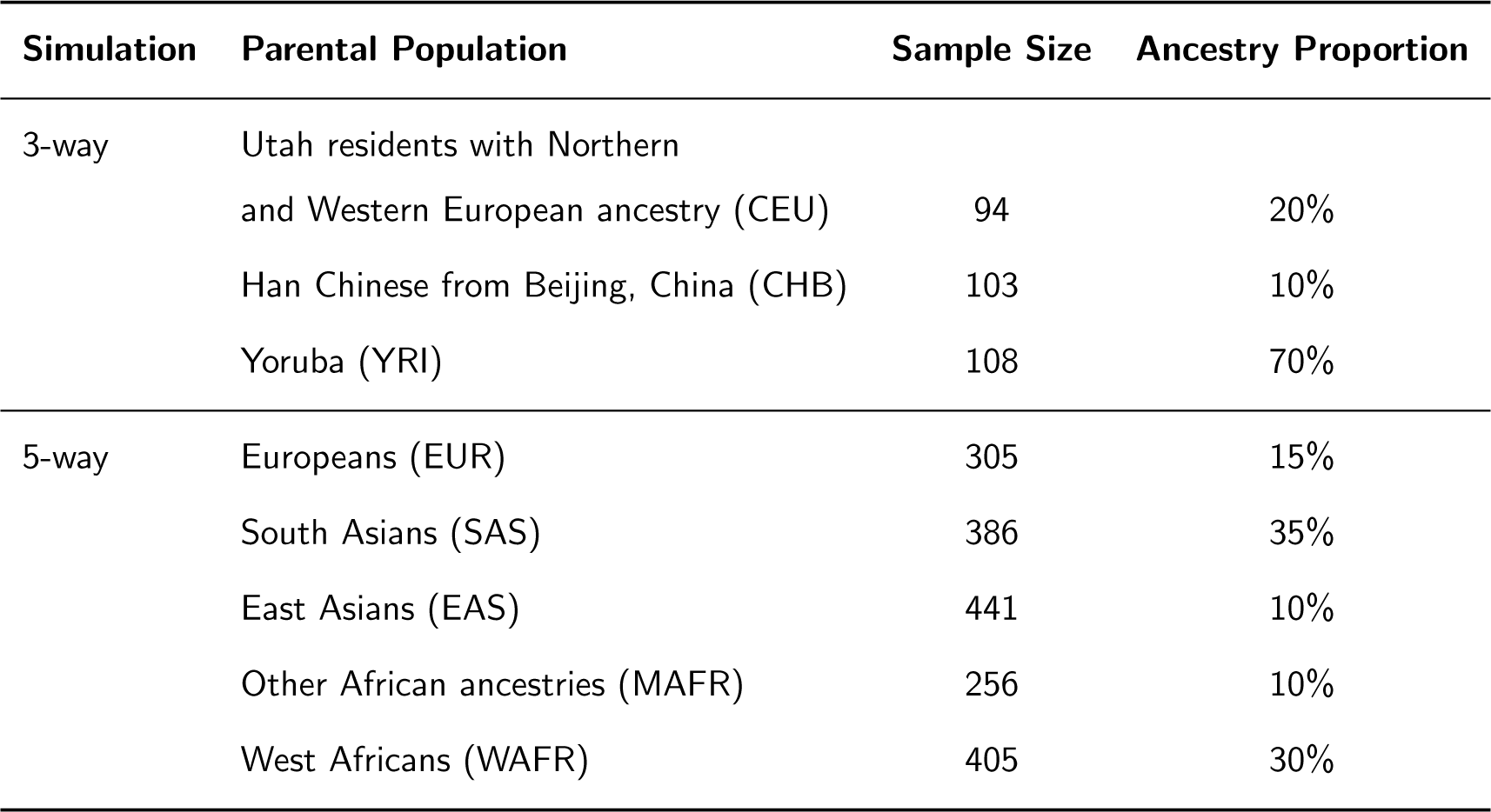
The table provides information on the parental reference populations for the 3-way and 5-way admixture simulations, their abbreviations, initial sample sizes, and the percentage of ancestry each population contributed in each scenario.

| Simulation | Parental Population | Sample Size | Ancestry Proportion |
| --- | --- | --- | --- |
| 3-way | Utah residents with Northern and Western European ancestry (CEU) | 94 | 20% |
|  | Han Chinese from Beijing, China (CHB) | 103 | 10% |
|  | Yoruba (YRI) | 108 | 70% |
| 5-way | Europeans (EUR) | 305 | 15% |
|  | South Asians (SAS) | 386 | 35% |
|  | East Asians (EAS) | 441 | 10% |
|  | Other African ancestries (MAFR) | 256 | 10% |
|  | West Africans (WAFR) | 405 | 30% |

In the 3-way simulation, we included 466,142 biallelic SNPs that were present in the 3 parental populations. We simulated four risk SNPs, where we selected one SNP each on chromosomes 2, 6, 11, and 15, and generated 2500 cases and 2500 controls. In the 5-way admixture scenario, we incorporated 623,330 biallelic SNPs that were present in all 5 parental populations and simulated 8 risk SNPs on chromosomes 2, 6, 11, 15, and 20. On chromosomes 2, 11, and 20, we selected two SNPs in each chromosome that were in high LD and one SNP each on chromosomes 6 and 15. In the 5-way scenario, however, we simulated two sets of datasets of different sample sizes: a dataset of 500 cases and 500 controls and another of 2500 cases and 2500 controls.

In the admixture simulation, we simulated different risk scenarios for the different chromosomes by varying the presence and strength of genotype risk on the risk variant simulated and the ancestry risk on the genomic region containing the variant. We simulated ancestry risk by simulating ancestry deviation between cases and controls in the region that contained the risk variants. In the 3-way simulation on chromosomes 2 and 11, we simulated strong genotype and ancestry risks; on chromosome 6, we simulated very strong ancestry risk and weak genotype risk; and on chromosome 15, we simulated weak genotype and ancestry risks. All the other chromosomes were simulated under a null model in this scenario. In the 5-way simulation, we simulated similar levels of risk in the 500 cases and 500 controls and 2,500 cases and 2,500 controls sample sizes. On chromosome 2, we simulated strong genotype and ancestry risks; on chromosomes 6 and 20, we simulated a strong genotype and no ancestry risk; on chromosomes 11 and 15, we simulated weak genotype and ancestry risks; and a null model on all the other chromosomes.

The risk SNPs simulated in the 3-way and 5-way scenarios and their respective homozygosity and heterozygisity relative risks specified for the cases are listed in **Table 4**. Depending on the MAF of the risk SNPs the specified risks introduced risk signals strength as indicated on **Table 5**.

**Table 4:**
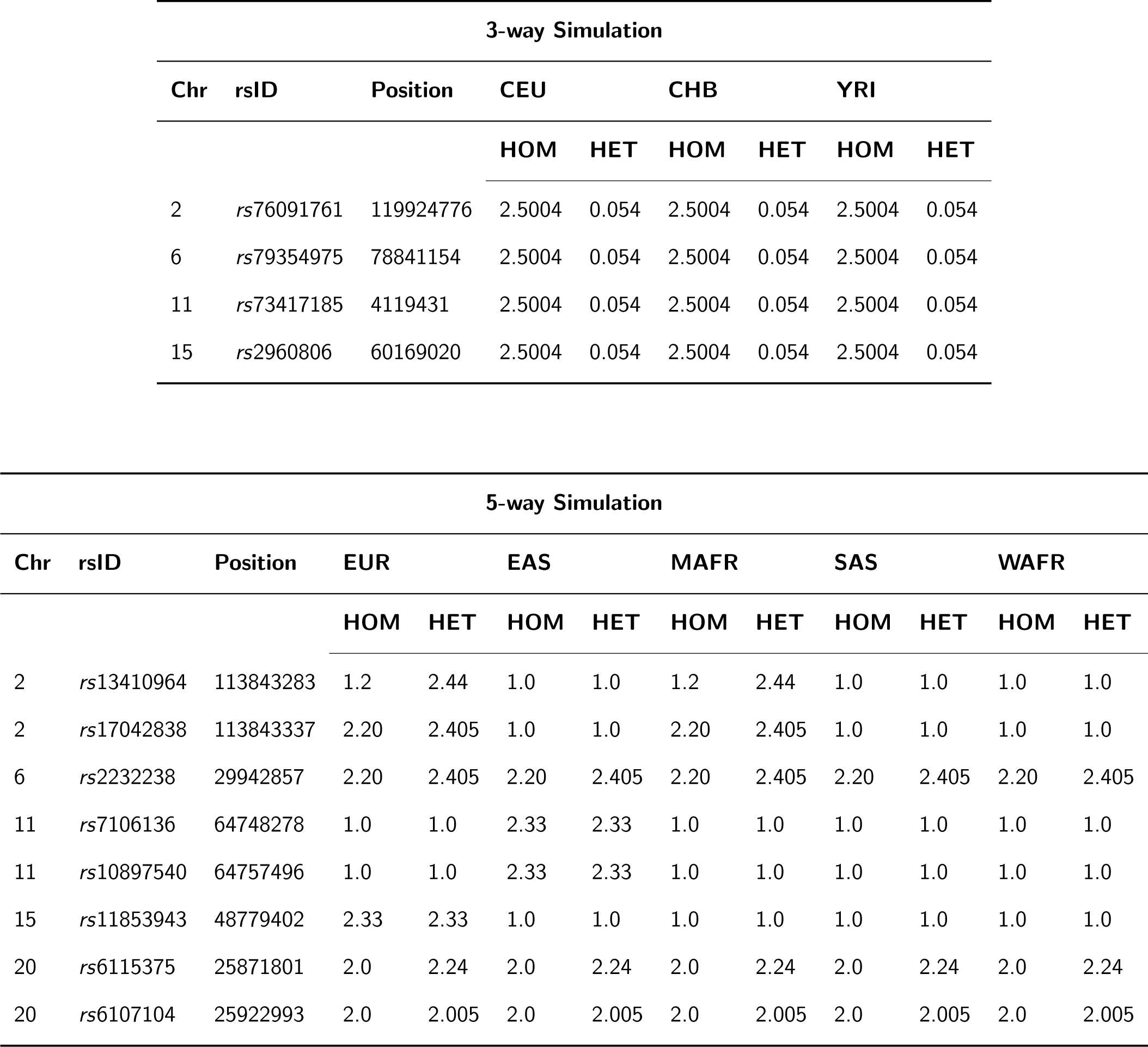
A list of simulated risk SNPs and the corresponding homozygosity (HOM) and heterozygosity (HET) relative risks specified during the isolated simulation of Europeans (EUR), East Asians (EAS), West Africans (WAFR), South Asians (SAS), and other African populations (MAFR) before the admixture process.

**Table 5:** The table lists the disease risk scenarios simulated in the 3-way and 5-way admixture simulations and the chromosomes containing the risk SNP. ✓ indicates a strong risk was simulated, (✓) indicates a weak risk was simulated, while ✗ indicates no risk was simulated.

| Simulation | Chromosome | Genotype Risk | Ancestry Risk |
| --- | --- | --- | --- |
| 3-way | 2 | ✓ | ✓ |
|  | 6 | ✓ | ✓ |
|  | 11 | (✓) | ✓ |
|  | 15 | (✓) | ✓ |
|  | Others | ✗ | ✗ |
| 5-way | 2 | ✓ | (✓) |
|  | 6 & 20 | ✓ | ✗ |
|  | 11 | (✓) | (✓) |
|  | 15 | (✓) | ✓ |
|  | Others | ✗ | ✗ |

### Assessment of Population Structure in the Simulated Datasets

We first assessed the structure of the simulated data for both the homogeneous and admixed populations. Since the simulation process was similar for the two sets of case-control datasets in the homogeneous populations and the 5-way admixture simulation, we used the 500 cases and 500 controls simulated datasets for this assessment. In the 3 populations, European, African, and admixed, we first merged the simulated GWAS datasets with their corresponding reference populations used in the simulation. We then obtained the first 10 PCs using principal component analysis (PCA) implemented in GCTA (Yang et al., 2011), and proceeded to plot the first and second PCs using the GENESIS (Buchmann and Hazelhurst, 2014) tool. We used two approaches to assess the global ancestry in the admixture simulation. We first ran the ADMIXTURE (Alexander et al., 2009) tool, using the supervised option, for the merged admixed datasets; then, secondly, we calculated the simulated global ancestry from the local ancestry block estimates generated by FractalSIM. We then plotted the two admixture tract plots for each scenario using the GENESIS tool.

Figures 2 and 3 show the PCA plots for the African and European population simulations, respectively, while Figures 4 and 6 are the PCA plots for the 3-way and 5-way admixed populations. The admixture tract plots for the 3-way and 5-way admixture simulations are shown in Figures 5 and 7, respectively.

**Figure 2:**
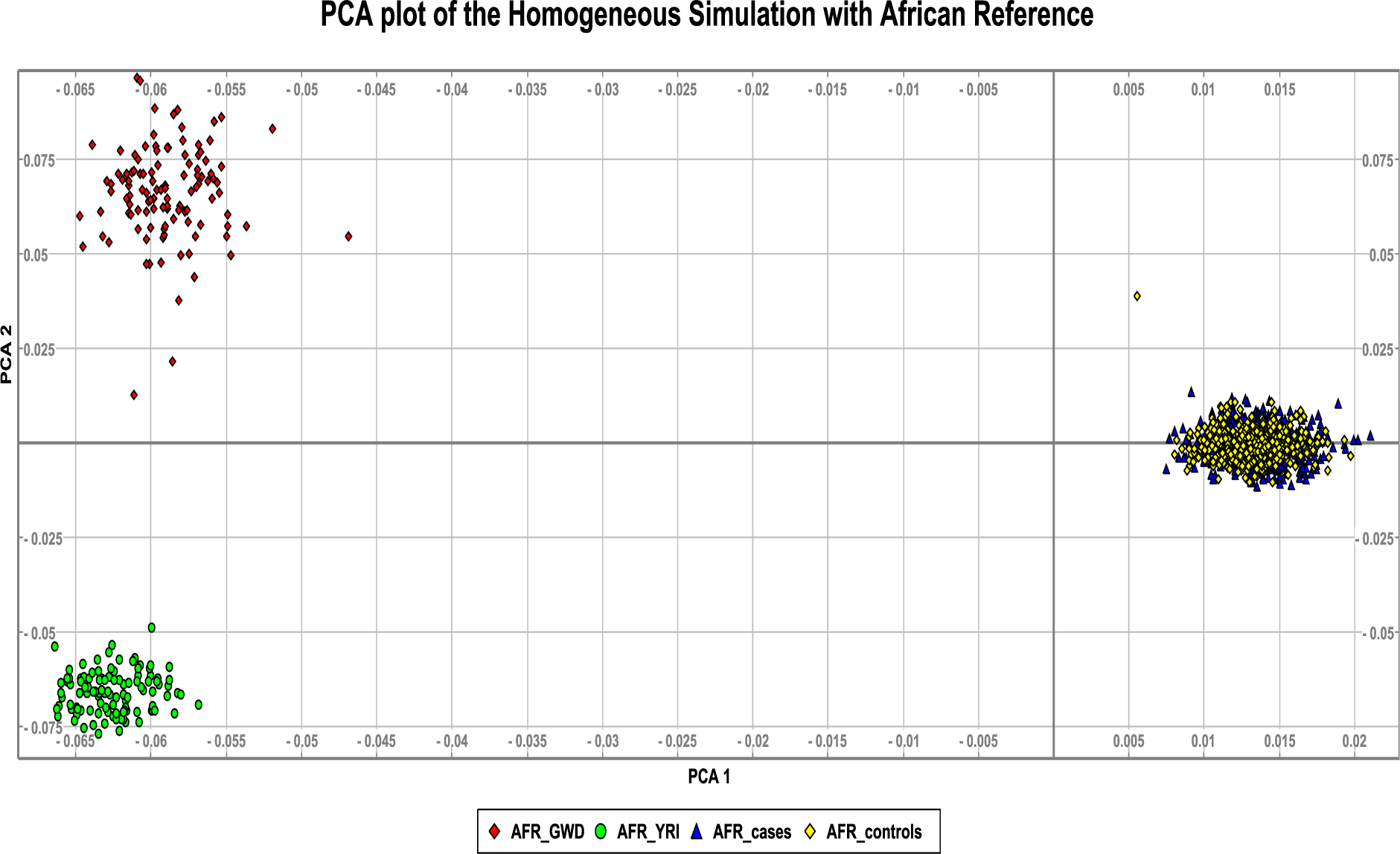
PCA plot of the simulated African population (500 cases and 500 controls) and the corre-sponding reference populations used in the simulation. AFR GWD indicates the Gambian Mandinka, AFR YRI indicates the Yoruba population, while AFR cases and AFR controls are the African simu-lated cases and controls.

**Figure 3:**
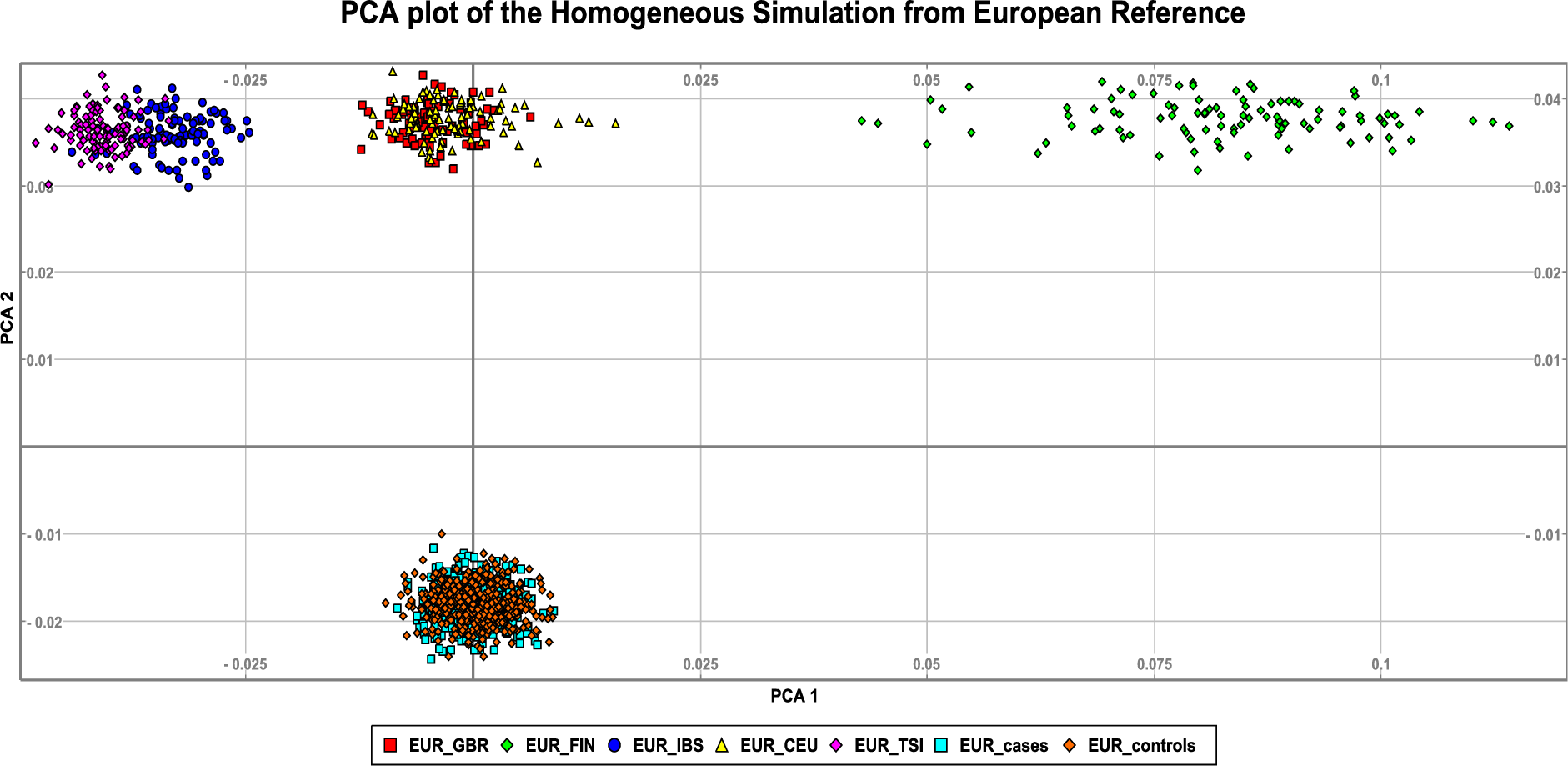
PCA plot of the simulated European population (500 cases and 500 controls) and the reference populations used in the simulation. EUR GBR are the British, EUR FIN are the Finnish, EUR IBS are the Iberian Spanish, EUR CEU are the Utah residents with Northern and Western European ancestry, and EUR TSI are the Toscani in Italy. The EUR cases and the EUR controls indicate the simulated European population.

**Figure 4:**
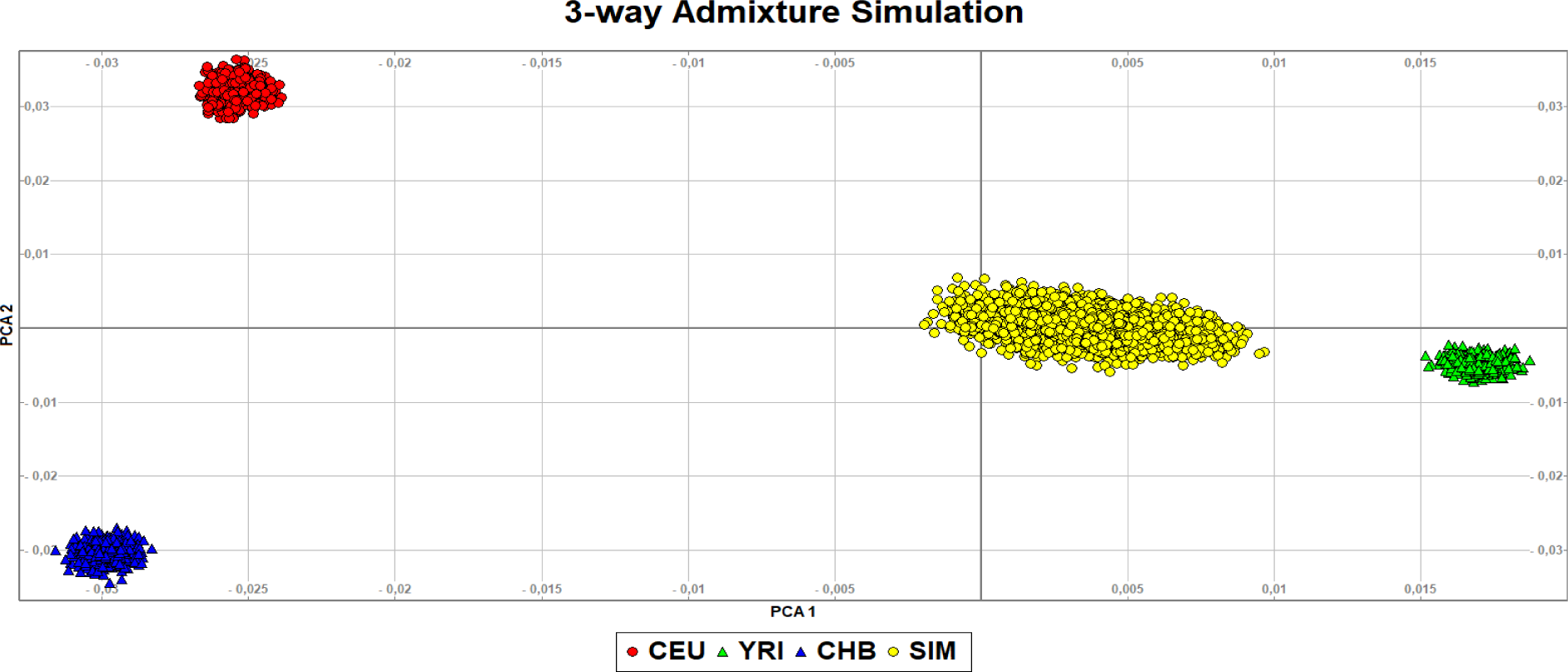
PCA plot of the 3-way admixture simulated population (2500 cases and 2500 controls) and the corresponding CEU, CHB, and YRI simulated in isolation from the reference populations. The simulated admixed population is labeled SIM.

**Figure 5:**
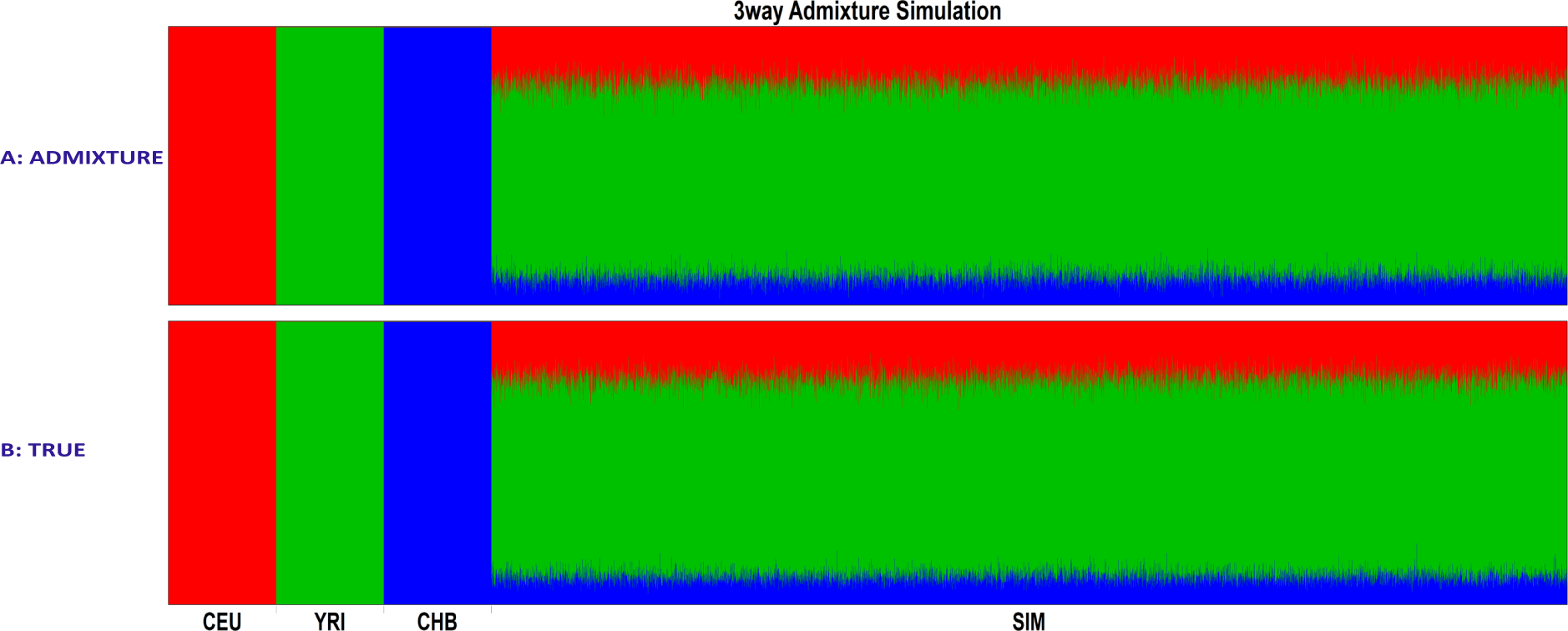
The admixture plot of the 3-way admixture simulated population (SIM), comprised of 2500cases and 2500 controls, and the corresponding CEU, YRI, and CHB reference populations used in the simulation. A: is the admixture block estimated by the ADMIXTURE tool and B: is the admixture block of the true simulated global ancestry proportion calculated from the ancestry block output by FractalSIM.

**Figure 6:**
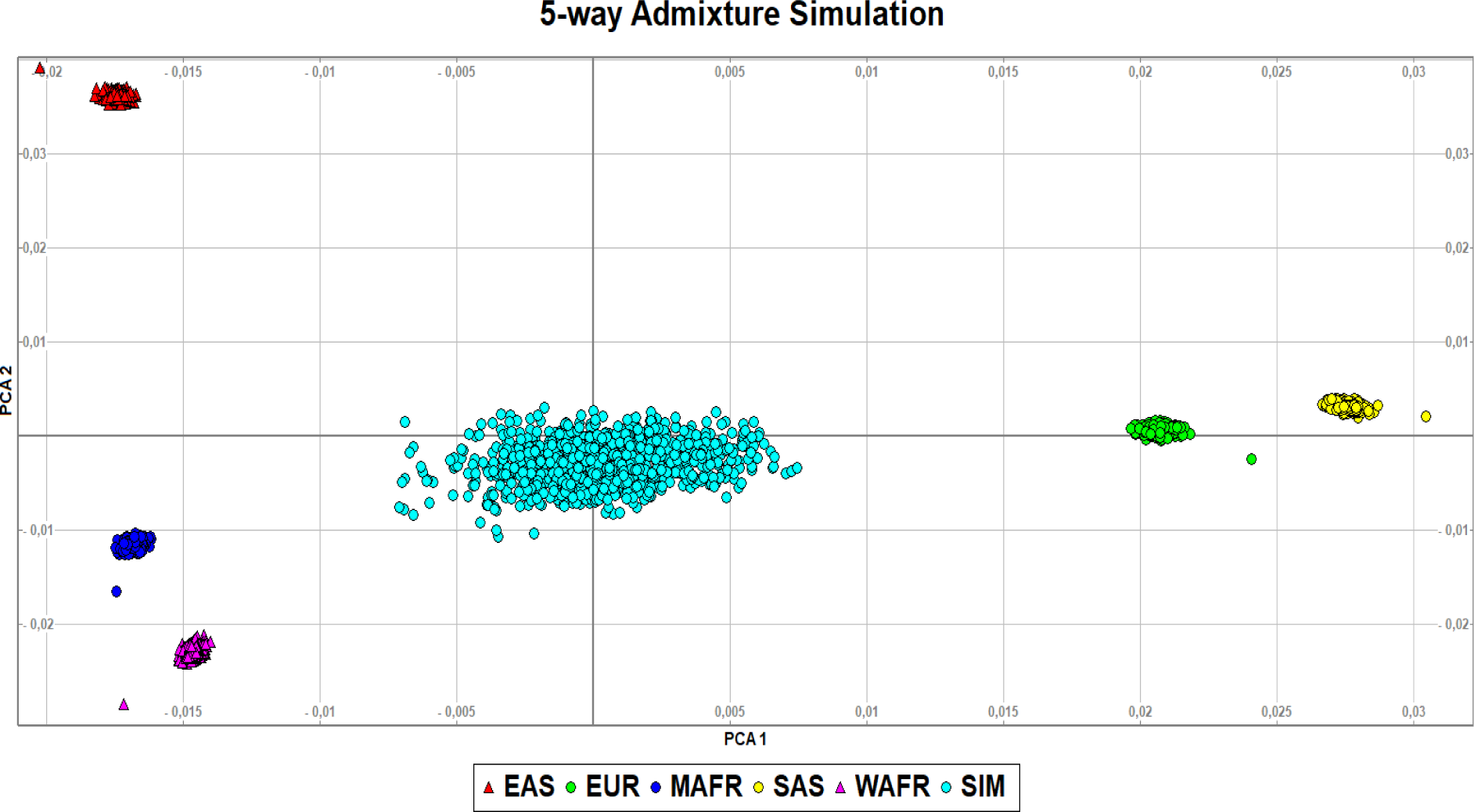
PCA plot of the 5-way admixed population simulation of 500 cases and 500 controls. ADM is the admixed group, while EAS, EUR, MAFR, SAS, and WAFR are the parental populations simulated in isolation from the reference ancestral populations.

**Figure 7:**
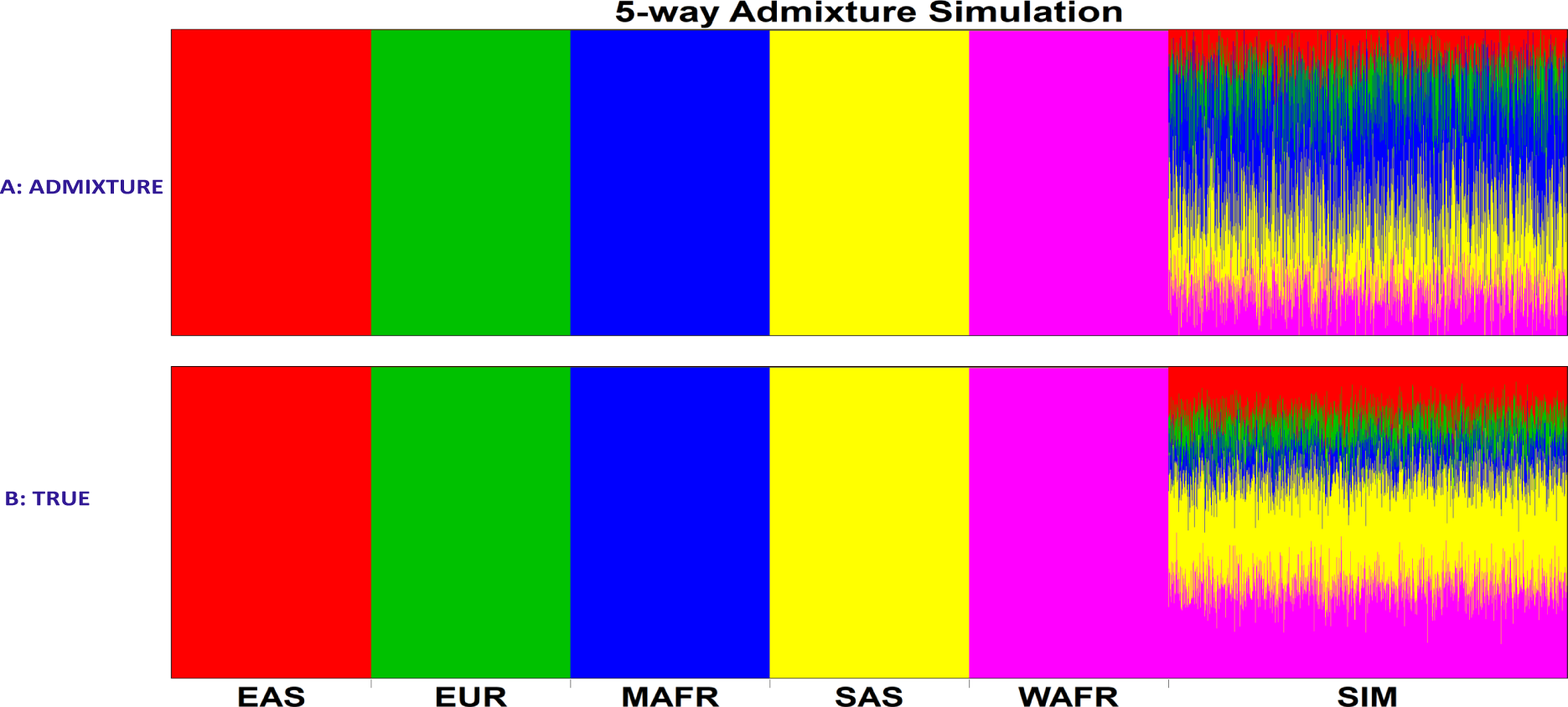
The admixture plot of the 5-way admixture simulation population (SIM), including 500 cases and 500 controls, and the respective EAS, EUR, MAFR, SAS, and WAFR parental populations. A: is the estimated admixture block produced by the ADMIXTURE tool, while B: is the estimated admixture block of the true simulated global ancestry proportion generated by FractalSIM.

On the PCA plots in Figures 2 and 3 we observed that the simulated African and European populations, for both the cases and controls, clustered together as would be expected in a homogenous population with no population structures. The simulated populations were also positioned between the merged reference populations on the PCA 2 axis for the African population and the PCA 1 axis for the European population. On the PCA 1 axis, the simulated African population was very close to the reference population by considering the range of the axis, and similarly, on the PCA 2 axis, the simulated European population was also very close to the reference population based on the range of the axis. This implies that the simulated cases and controls were genetically close to the respective merged African and European reference samples.

For the admixed population, we observed on the PCA plots in Figures 4 and 6 that the admixed samples were confined within their respective reference parental populations, for both the 3-way and 5-way populations. We also observed that the simulated population was spread out, as would be expected for an admixed population. The simulated 3-way admixed population was closer to the YRI population, which contributed 70% of the ancestry, while the 5-way admixed population is spread out further away from the EAS population but closer to the MAFR and SAS populations, which contributed larger proportions of the ancestry. For the admixture tract, for both the 3-way and 5-way scenarios, in Figures 5 and 7 we observe that the ADMIXTURE tool estimates the global ancestry close to the true estimates but performs better in a 3-way simulation than a 5-way simulation.

The PCA and admixture plots indicate that the structure of the simulated populations met the criteria of the population that we required for the downstream analysis.

### Association Analysis of the Simulated Populations

In this endeavor, we investigated five commonly used GWAS tools for both homogeneous and multi-way admixed populations using the simulated GWAS datasets described above. We included LMM-based approaches EMMAX (Kang et al., 2010), GCTA (Yang et al., 2011) and GEMMA (Zhou and Stephens, 2012), as well as the most widely used GWAS analysis tools PLINK (Purcell et al., 2007), and SNPTEST (Marchini and Howie, 2010). For the homogeneous African and European population we considered the standard PLINK association and under a logistic model that allowed us to include covariates, which we labeled PLINK-Logistic. For the admixture simulations we only considered PLINK-Logistic. For GCTA, we considered two association approaches included in the tool. In the first approach, the GRM used includes the chromosome with the SNP being tested for association, which we labeled GCTA, while the second approach uses a GRM that excludes the chromosome that contains the SNP being tested for association, which we label GCTA-LOCO (leave one chromosome out). This approach is an extension of GCTA to eliminate proximal contamination that may be introduced in the association analysis when this chromosome is included in the calculation of the GRM. Similarly, in SNPTEST, we considered both the frequentist association approach (which we refer to as SNPTEST-Frequentist) and the Bayesian approach (which we refer to as SNPTEST-Bayesian).

We first obtained the first 10 PCs under each simulated set of data using GCTA. For the homogeneous populations, we included 5 PCs as covariates when running PLINK-Logistic and SNPTEST, as based on the PCA plots, we did not observe structures in the homogeneous cohorts. In the admixture populations, however, we included 10 PCs as covariates in the association test to control for global ancestry. No missingness was observed in the datasets, and all the simulated samples were retained for the association analysis. We considered only common SNPs when running the association tests.

We thus ran the association analysis using eight disease-scoring statistics for the homogeneous population and seven for the admixed population. We then obtained the corresponding summary statistics and Manhattan plots. To correct for multiple tests, we used the Bonferroni correction approach. Since the number of SNPs in our homogeneous population was *>* 1, 000, 000, we used a standard genome-wide significance of 1.0 *×* 10*^−^*^08^ for all the frequentist tests. The significance threshold for the 3-way admixed population was 1.576984 *×* 10*^−^*^07^; for the 5-way admixed population, for the smaller sample size, 8.480081 *×* 10*^−^*^08^; and for the larger sample size, 8.479046 *×* 10 *−* 08. We used *log*(BF) of 4.61 as the significant threshold for the BF factor for the SNPTEST-Bayesian test, using Jeffrey’s scale of evidence (Jeffreys, 1961).

## Results

### Assessment of the European and African Simulation GWAS Analysis

The Manhattan plots for the homogeneous European and African populations are shown in Figures 8 to 11 and the corresponding summary statistics tables for the simulated risk SNPs are on **Tables 6** to **21**. In both simulations, we observed that, for all the tools assessed with the small sample size, none detected the signal on chromosome 11. However, for the European population, the LMM-based tools, GEMMA, GCTA, and GCTA-LOCO, capture the signals on 4 of the chromosomes, while PLINK and PLINK-Logistic detect significant signals on 3 of the chromosomes. Though EMMAX and SNPTEST detect 3 out of the 5 simulated risk regions at this sample size for the European population, they eliminate the risk SNP on chromosome 6 from the analysis as part of internal quality controls, and thus no significant SNP was observed. In comparison, in the African population with the smaller sample size, we observed that all the tools were only able to capture the signals on chromosomes 2 and 6 at significant levels, and the signals on chromosomes 15 and 20 only at marginal significance thresholds.

**Figure 8:**
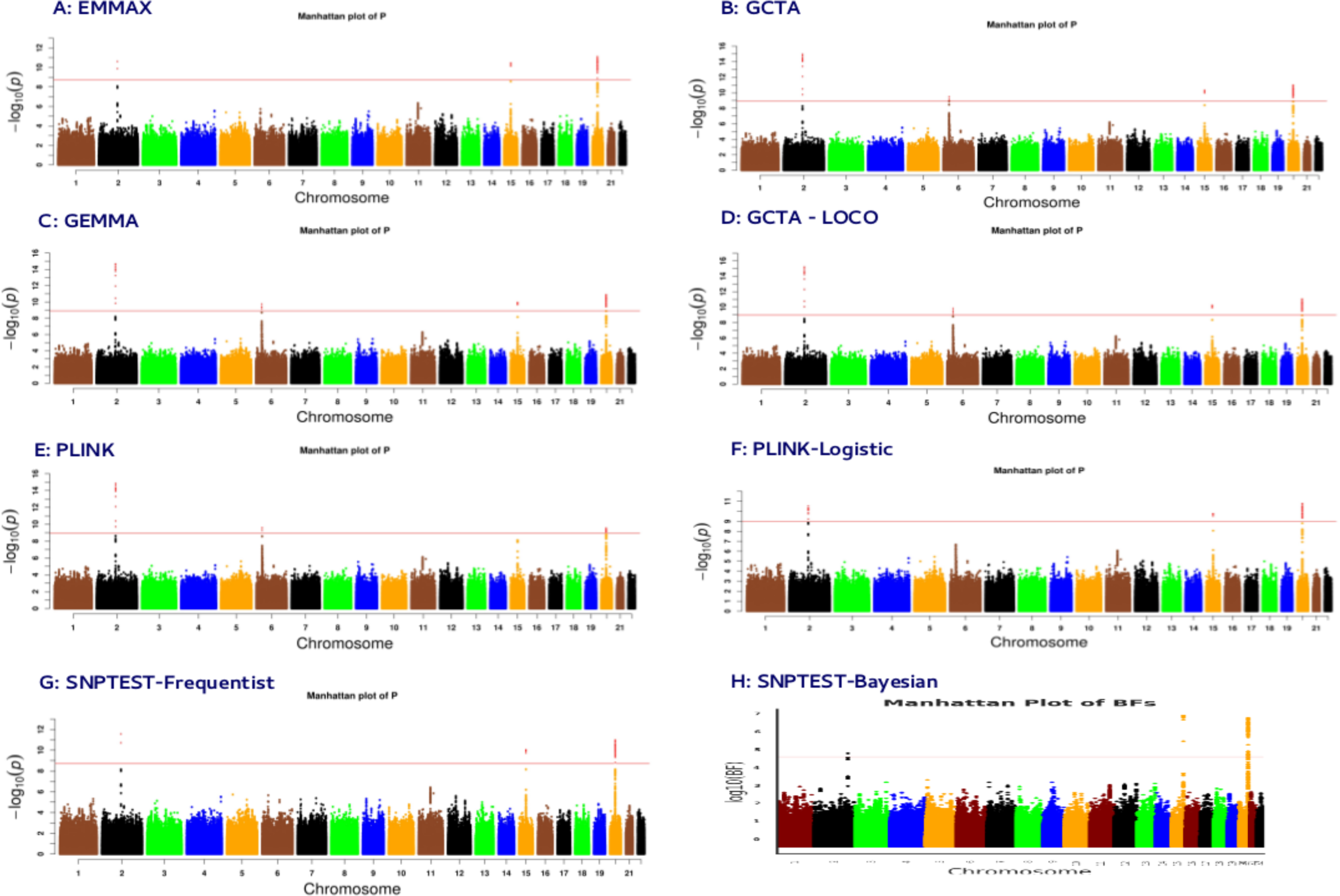
Manhattan plot of the GWAS summary statistics of the homogeneous simulation of 500 cases and 500 controls from the European reference population for 8 disease scoring statistics, corresponding to 5 tools. The red line indicates the GWAS significance level.

**Figure 9:**
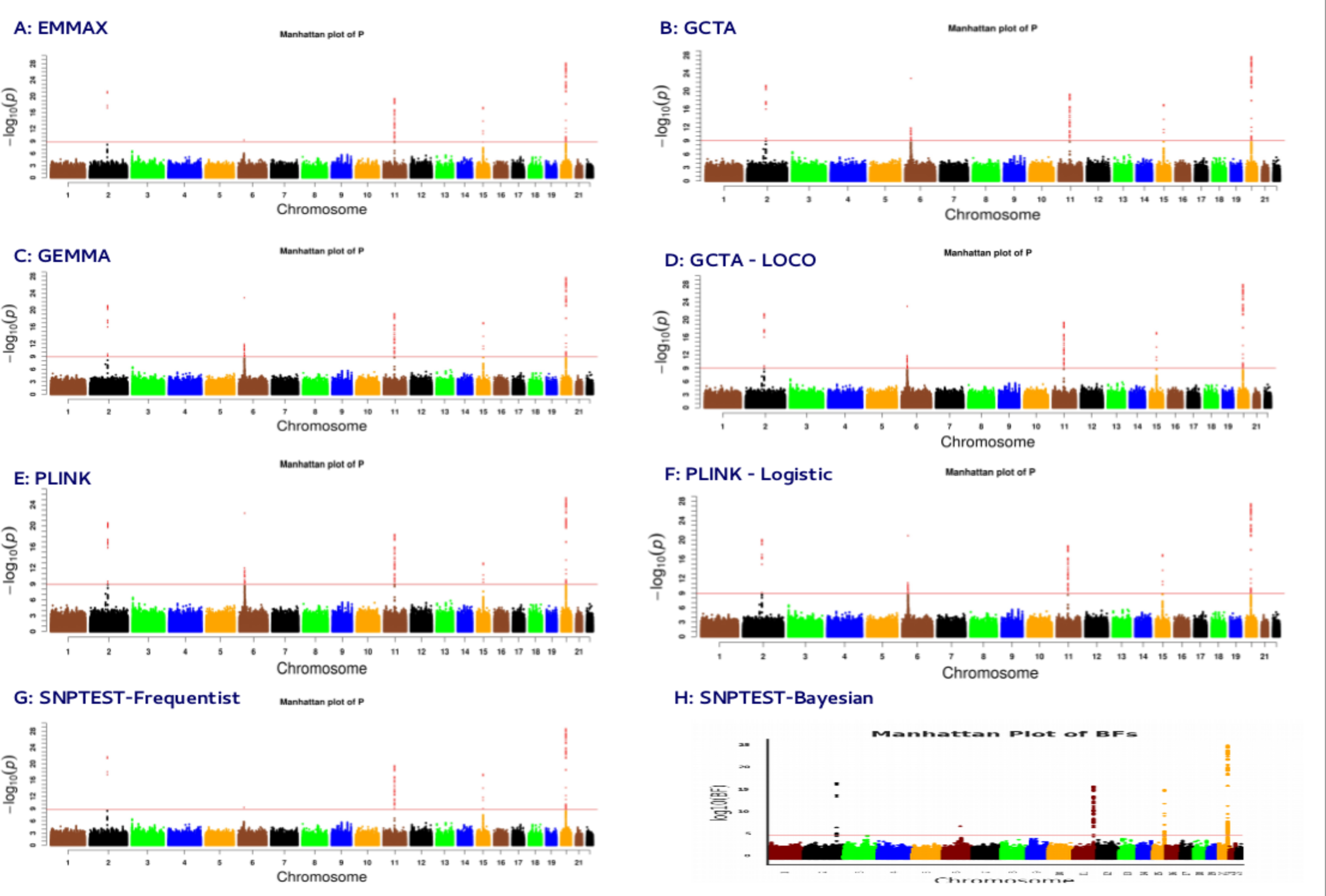
Manhattan plot of the GWAS summary statistics of the homogeneous simulation of 2500 cases and 2500 controls from the European reference population for 8 disease scoring statistics, corresponding to 5 tools. The red line indicates the GWAS significance level.

**Figure 10:**
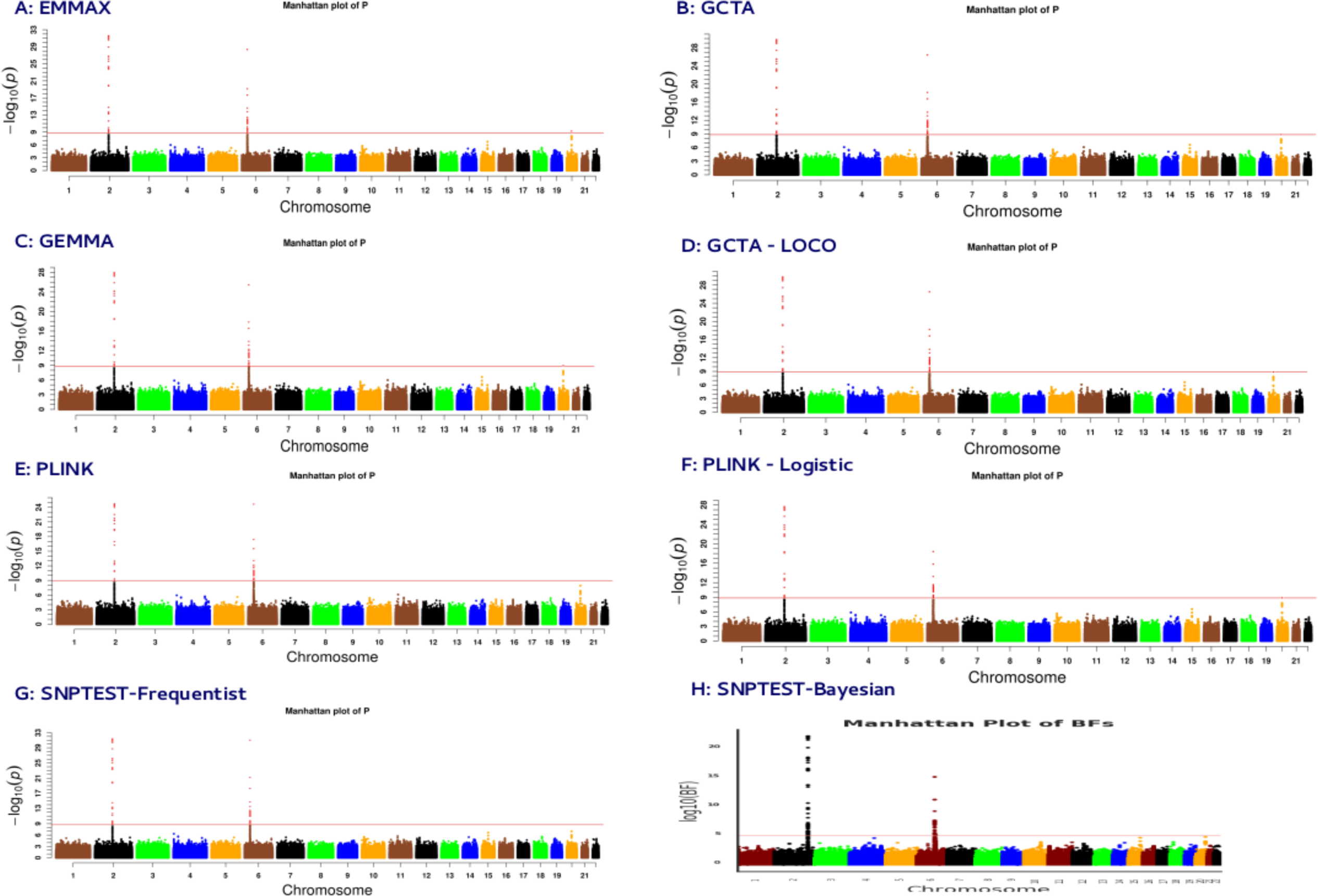
Manhattan plot of the GWAS summary statistics of the homogeneous simulation of 500 cases and 500 controls from the African reference population for 8 disease scoring statistics, corresponding to 5 tools. The red line indicates the GWAS significance level.

**Figure 11:**
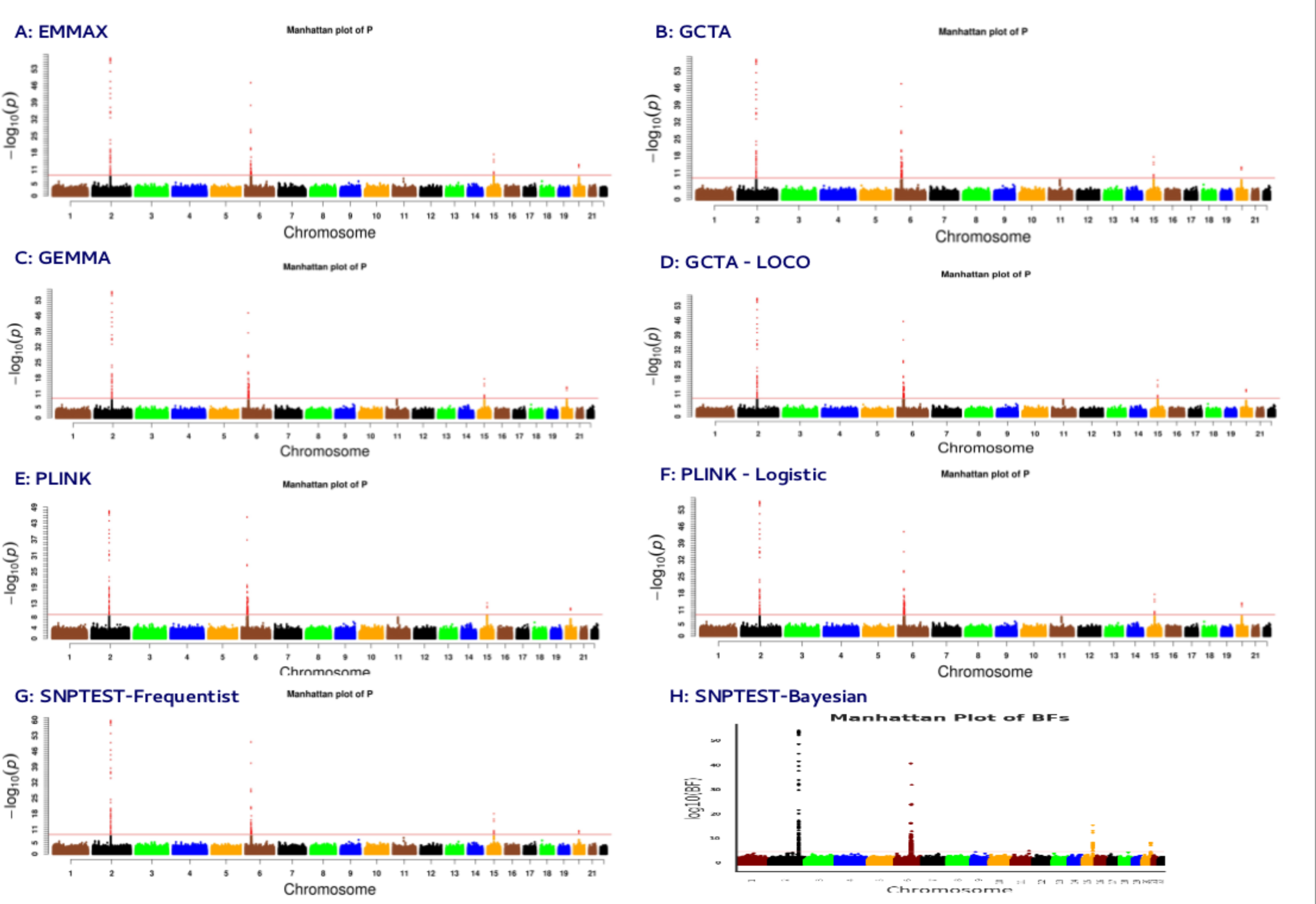
Manhattan plot of the GWAS summary statistics of the homogeneous simulation of 2500 cases and 2500 controls from the African reference population for 8 disease scoring statistics, corresponding to 5 tools. The red line indicates the GWAS significance level.

**Table 6:**
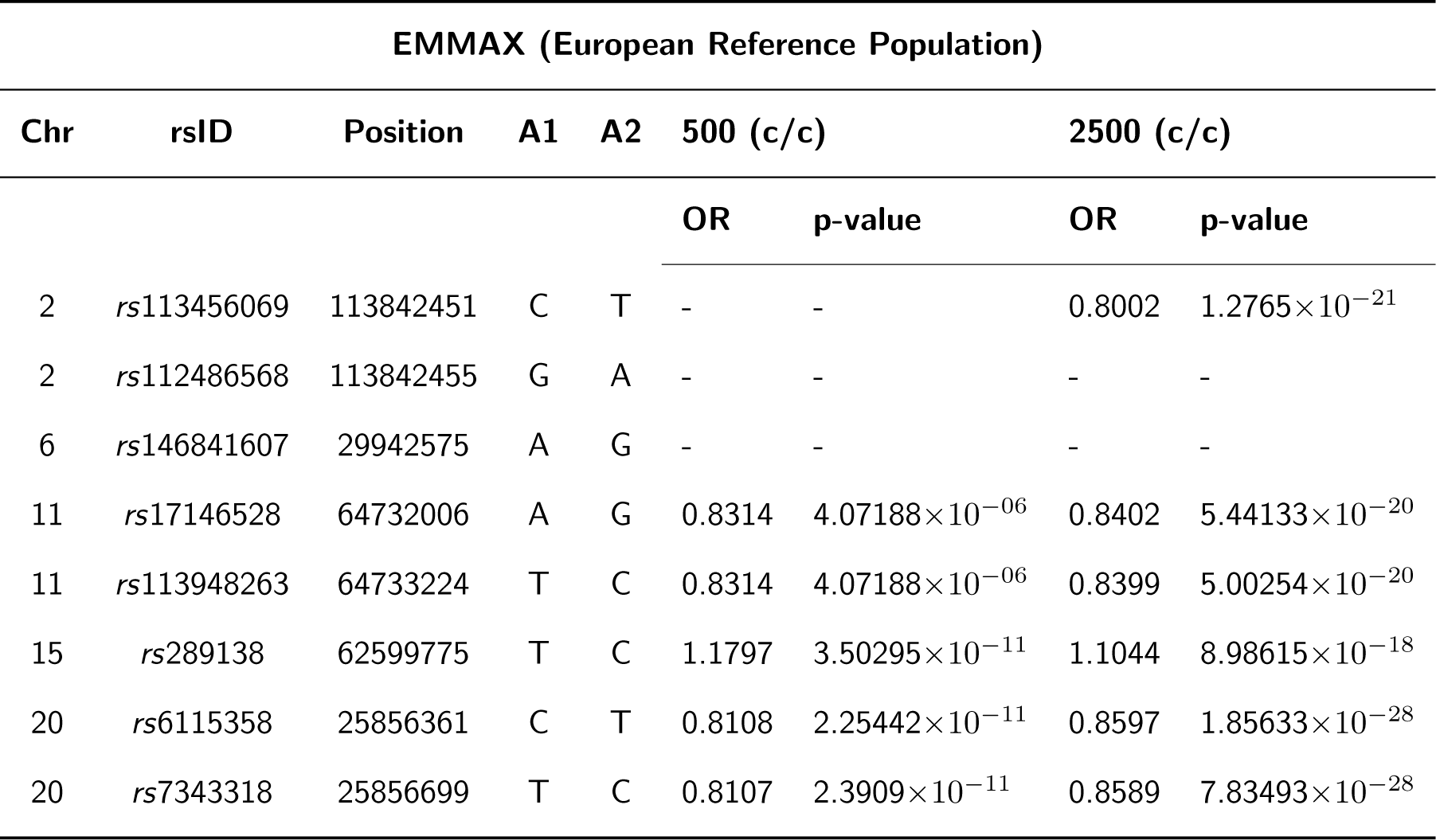
GWAS summary statistics from EMMAX of the simulated risk SNPs in the homogeneous simulation of 500 and 2500 samples from a European reference population of 489 samples. **x**(**c***/***c**) refers to the **x** number of cases and controls.

**Table 7:**
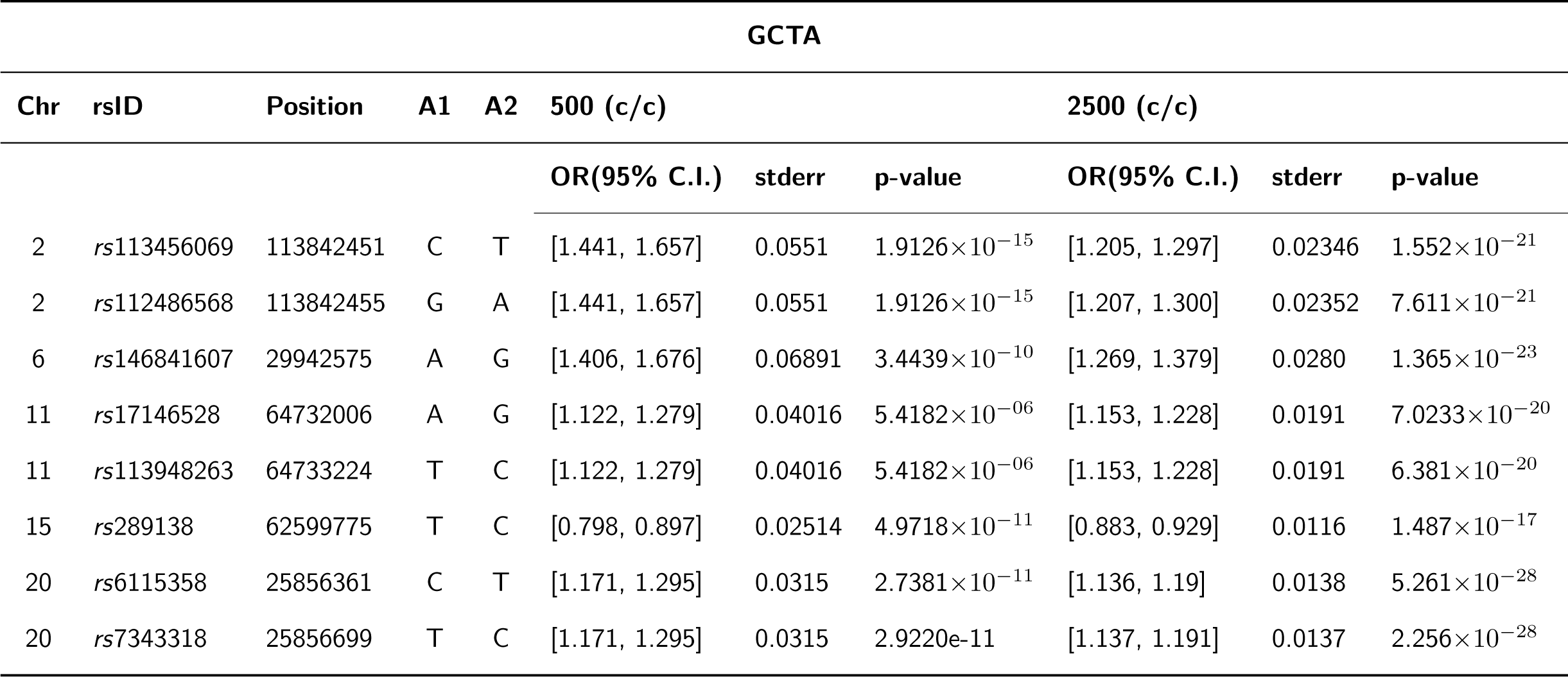
GWAS summary statistics from GCTA of the simulated risk SNPs in the homogeneous simulation of 500 and 2500 samples from a European reference population of 489 samples. **x**(**c***/***c**) refers to the **x** number of cases and controls.

| GCTA |  |  |  |  |  |  |  |  |  |  |
| --- | --- | --- | --- | --- | --- | --- | --- | --- | --- | --- |
| Chr | rsID | Position | A1 | A2 | 500 (c/c) |  |  | 2500 (c/c) |  |  |
|  |  |  |  |  | OR(95% C.I.) | stderr | p-value | OR(95% C.I.) | stderr | p-value |
| 2 | rs113456069 | 113842451 | C | T | [1.441, 1.657] | 0.0551 | 1.9126×10<sup>-15</sup> | [1.205, 1.297] | 0.02346 | 1.552×10<sup>-21</sup> |
| 2 | rs112486568 | 113842455 | G | A | [1.441, 1.657] | 0.0551 | 1.9126×10<sup>-15</sup> | [1.207, 1.300] | 0.02352 | 7.611×10<sup>-21</sup> |
| 6 | rs146841607 | 29942575 | A | G | [1.406, 1.676] | 0.06891 | 3.4439×10<sup>-10</sup> | [1.269, 1.379] | 0.0280 | 1.365×10<sup>-23</sup> |
| 11 | rs17146528 | 64732006 | A | G | [1.122, 1.279] | 0.04016 | 5.4182×10<sup>-06</sup> | [1.153, 1.228] | 0.0191 | 7.0233×10<sup>-20</sup> |
| 11 | rs113948263 | 64733224 | T | C | [1.122, 1.279] | 0.04016 | 5.4182×10<sup>-06</sup> | [1.153, 1.228] | 0.0191 | 6.381×10<sup>-20</sup> |
| 15 | rs289138 | 62599775 | T | C | [0.798, 0.897] | 0.02514 | 4.9718×10<sup>-11</sup> | [0.883, 0.929] | 0.0116 | 1.487×10<sup>-17</sup> |
| 20 | rs6115358 | 25856361 | C | T | [1.171, 1.295] | 0.0315 | 2.7381×10<sup>-11</sup> | [1.136, 1.19] | 0.0138 | 5.261×10<sup>-28</sup> |
| 20 | rs7343318 | 25856699 | T | C | [1.171, 1.295] | 0.0315 | 2.9220e-11 | [1.137, 1.191] | 0.0137 | 2.256×10<sup>-28</sup> |

**Table 8:**
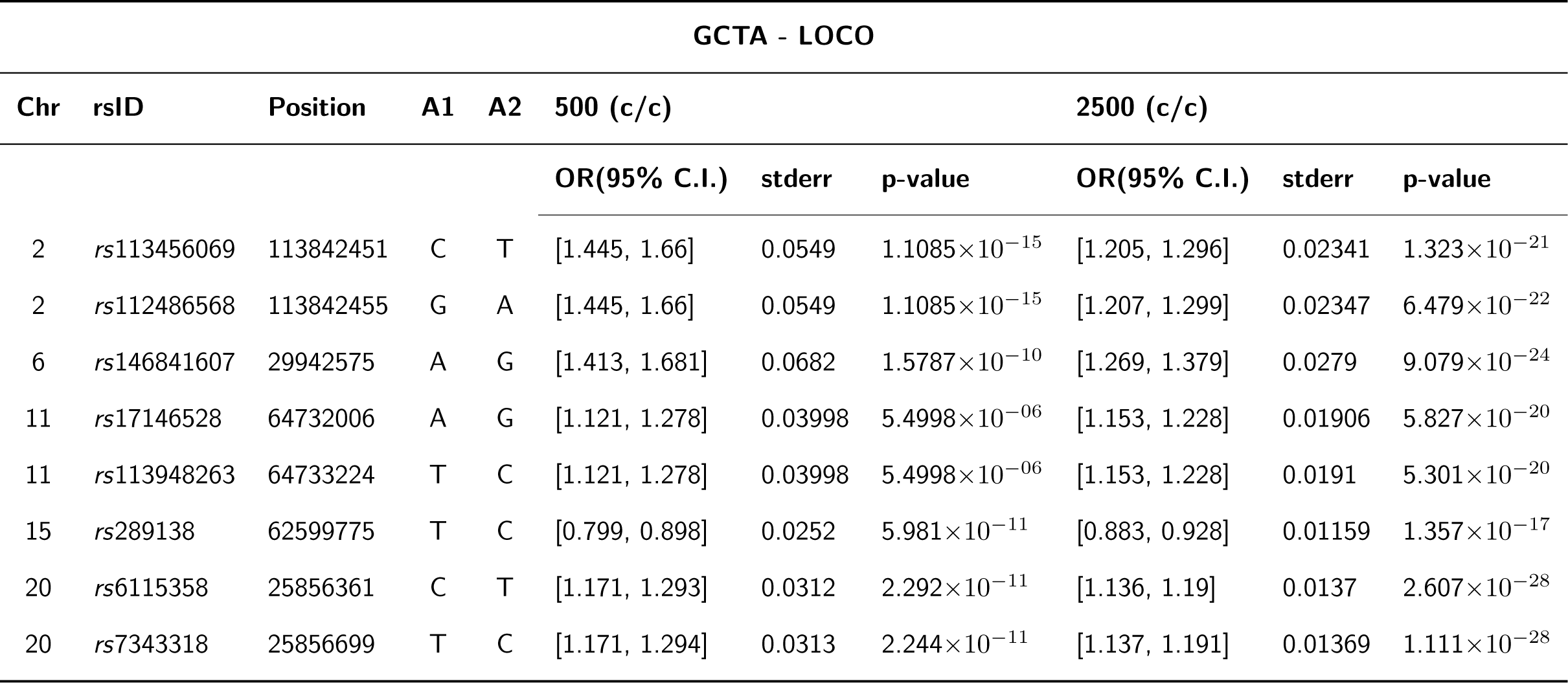
GWAS summary statistics from GCTA - LOCO of the simulated risk SNPs in the homogeneous simulation of 500 and 2500 samples from a European reference population of 489 samples. **x**(**c***/***c**) refers to the **x** number of cases and controls.

**Table 9:**
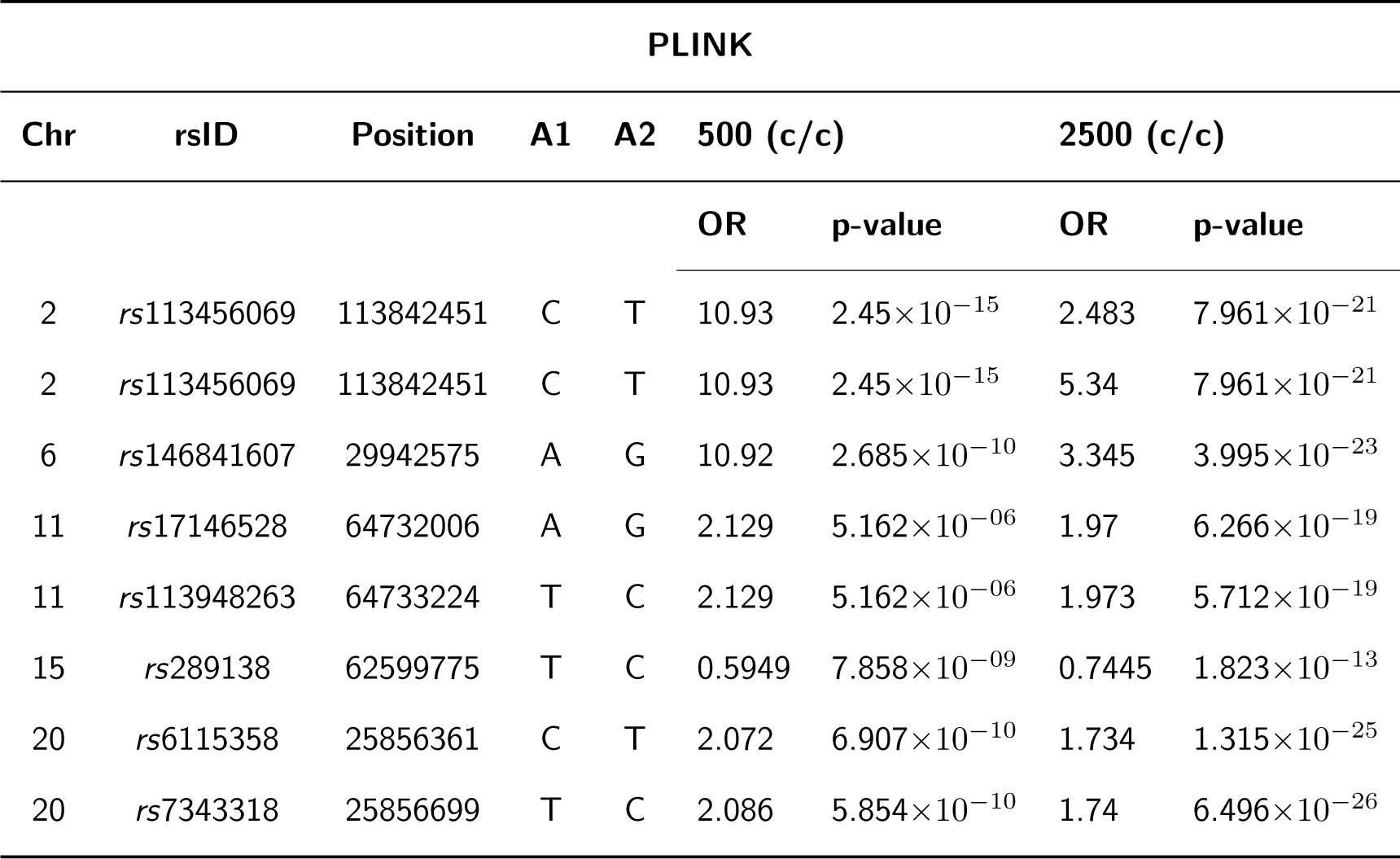
GWAS summary statistics from PLINK of the simulated risk SNPs in the homogeneous simulation of 500 and 2500 samples from a European reference population of 489 samples. **x**(**c***/***c**) refers to the **x** number of cases and controls.

**Table 10:**
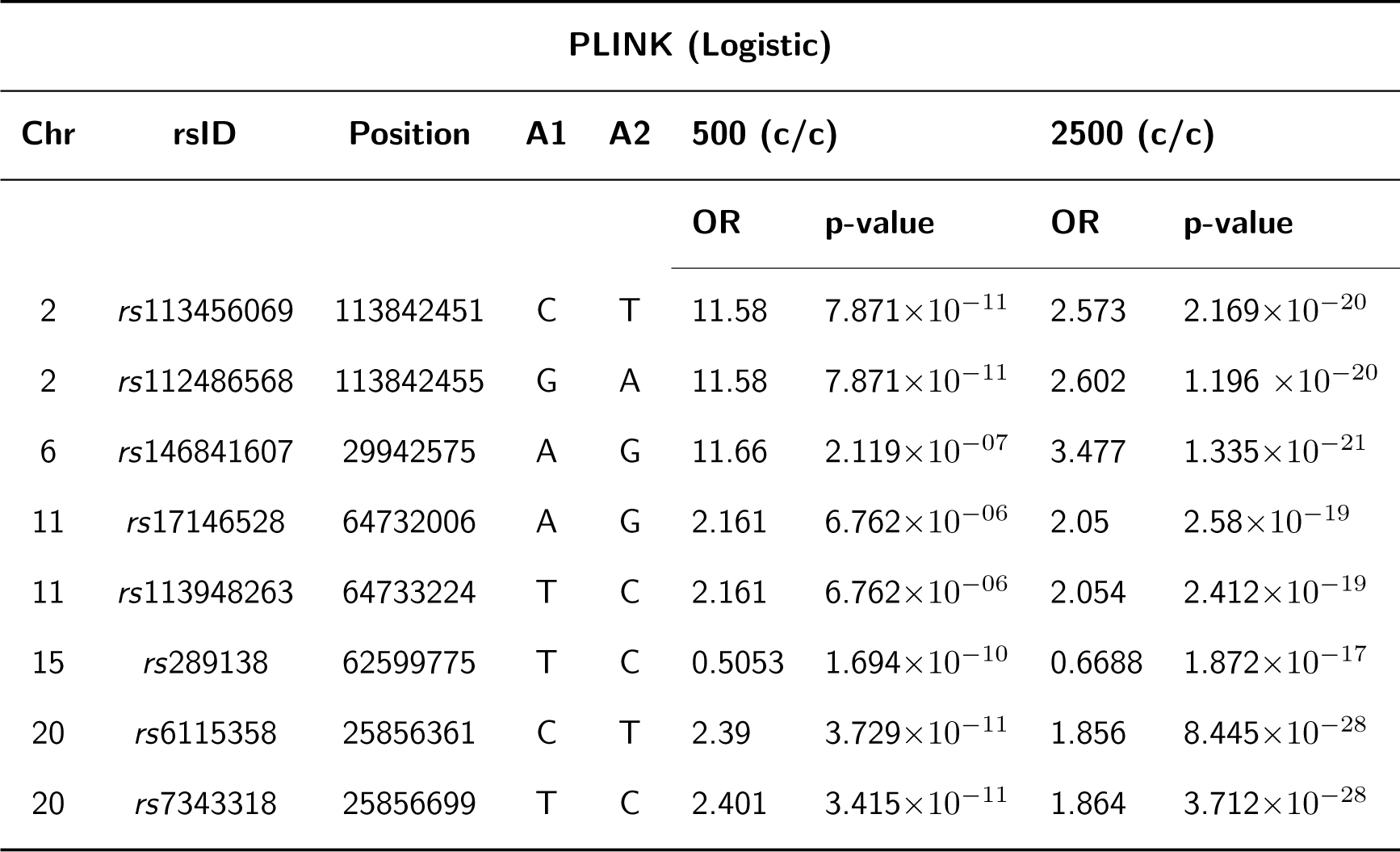
GWAS summary statistics from PLINK (Logistic) of the simulated risk SNPs in the homogeneous simulation of 500 and 2500 samples from a European reference population of 489 samples. **x**(**c***/***c**) refers to the **x** number of cases and controls.

**Table 11:**
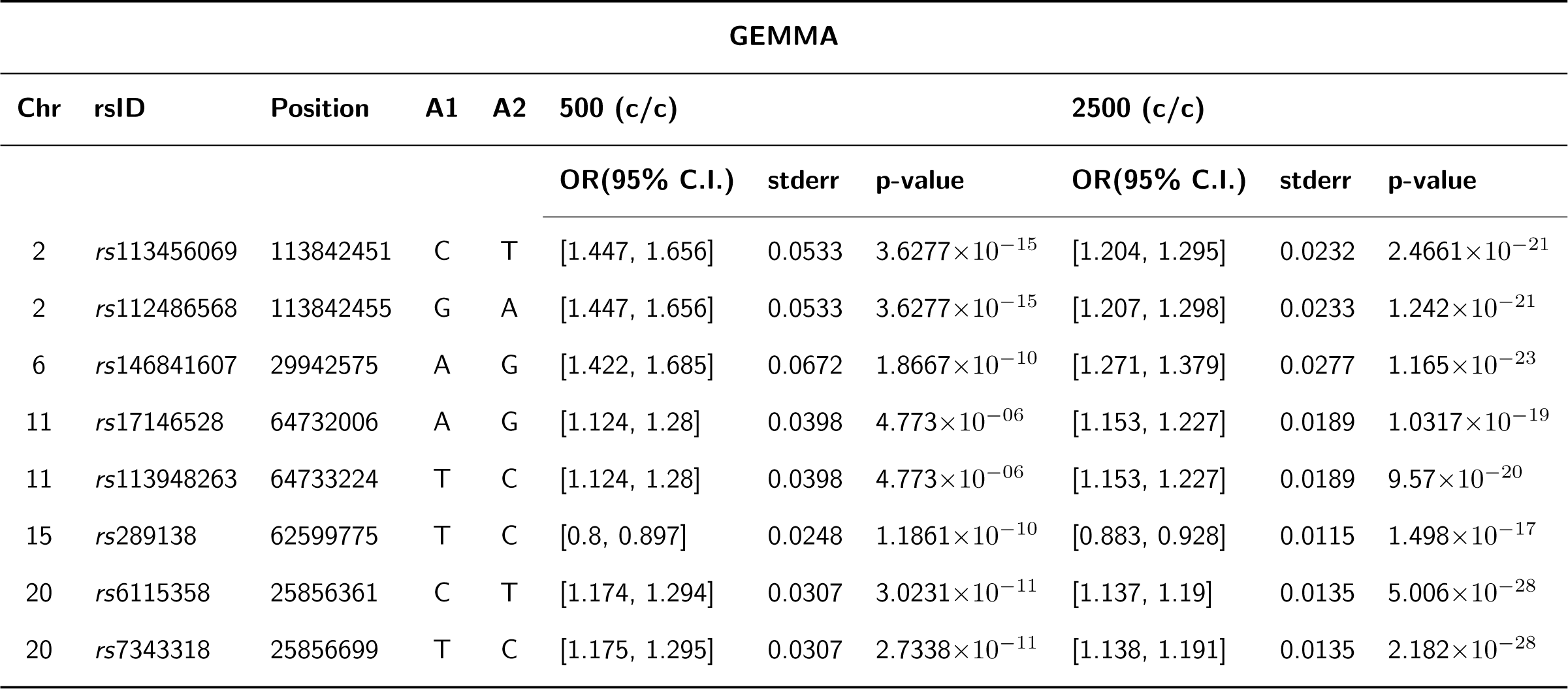
GWAS summary statistics from GEMMA of the simulated risk SNPs in the homogeneous simulation of 500 and 2500 samples from a European reference population of 489 samples. **x**(**c***/***c**) refers to the **x** number of cases and controls.

**Table 12:**
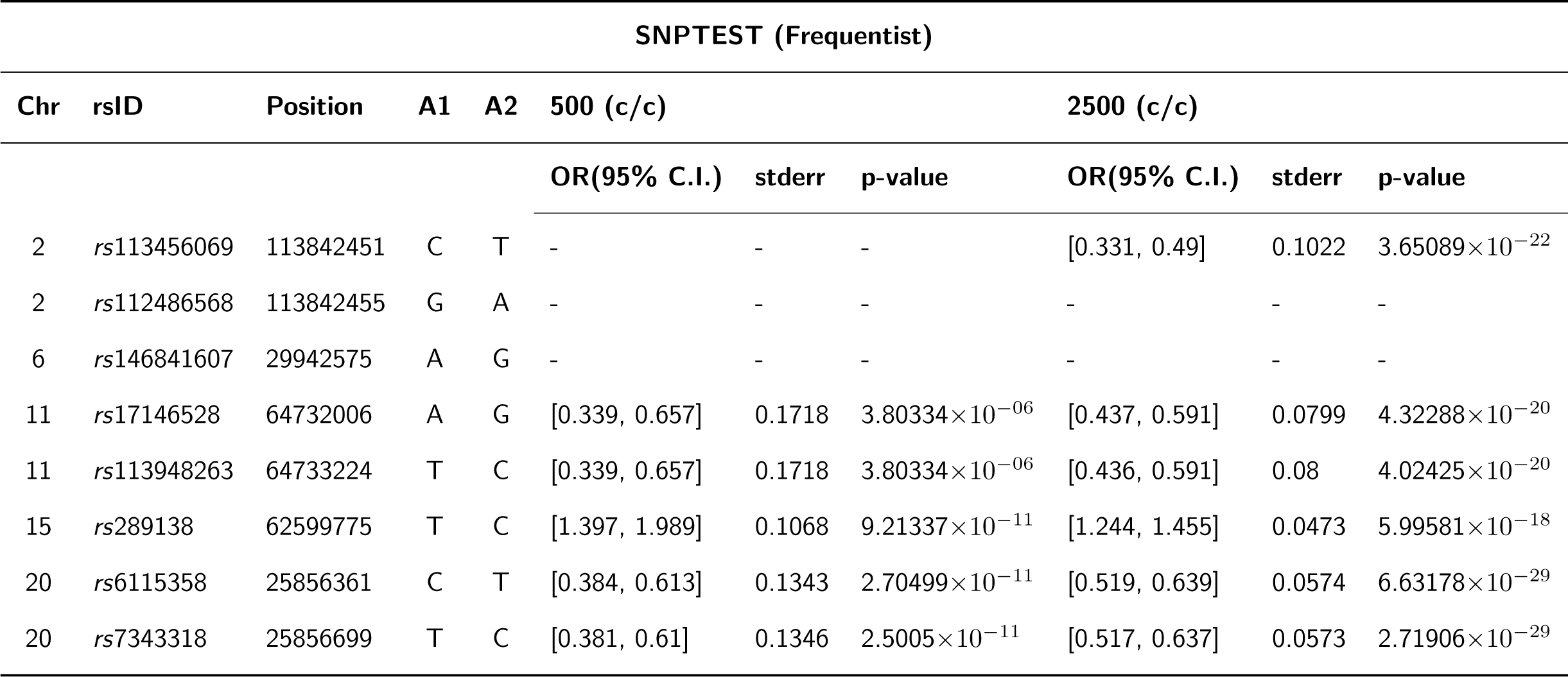
GWAS summary statistics from SNPTEST (Frequentist) of the simulated risk SNPs in the homogeneous simulation of 500 and 2500 samples from a European reference population of 489 samples. **x**(**c***/***c**) refers to the **x** number of cases and controls.

**Table 13:**
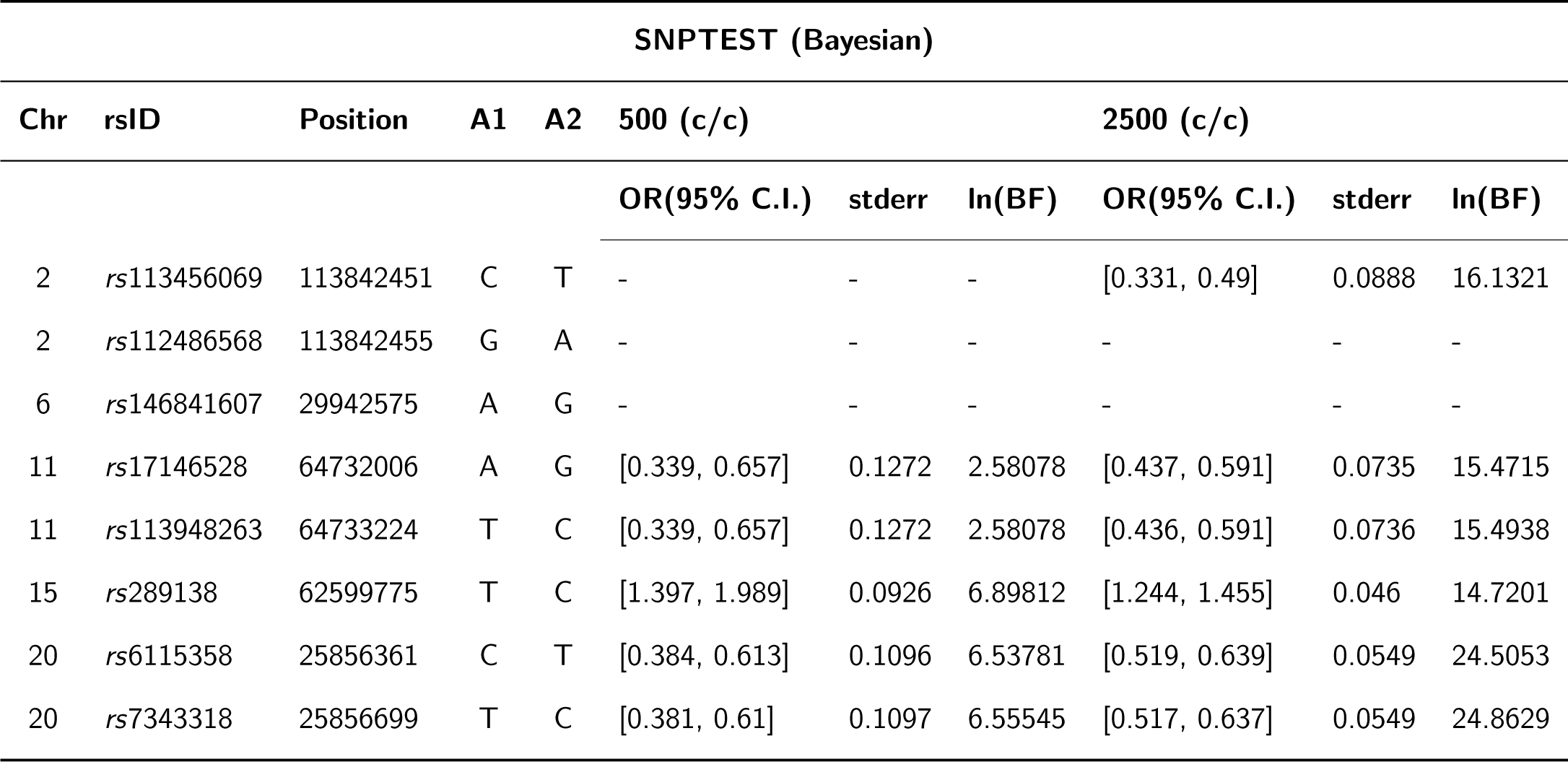
GWAS summary statistics from SNPTEST (Bayesian) of the simulated risk SNPs in the homogeneous simulation of 500 and 2500 samples from a European reference population of 489 samples. **x**(**c***/***c**) refers to the **x** number of cases and controls.

**Table 14:**
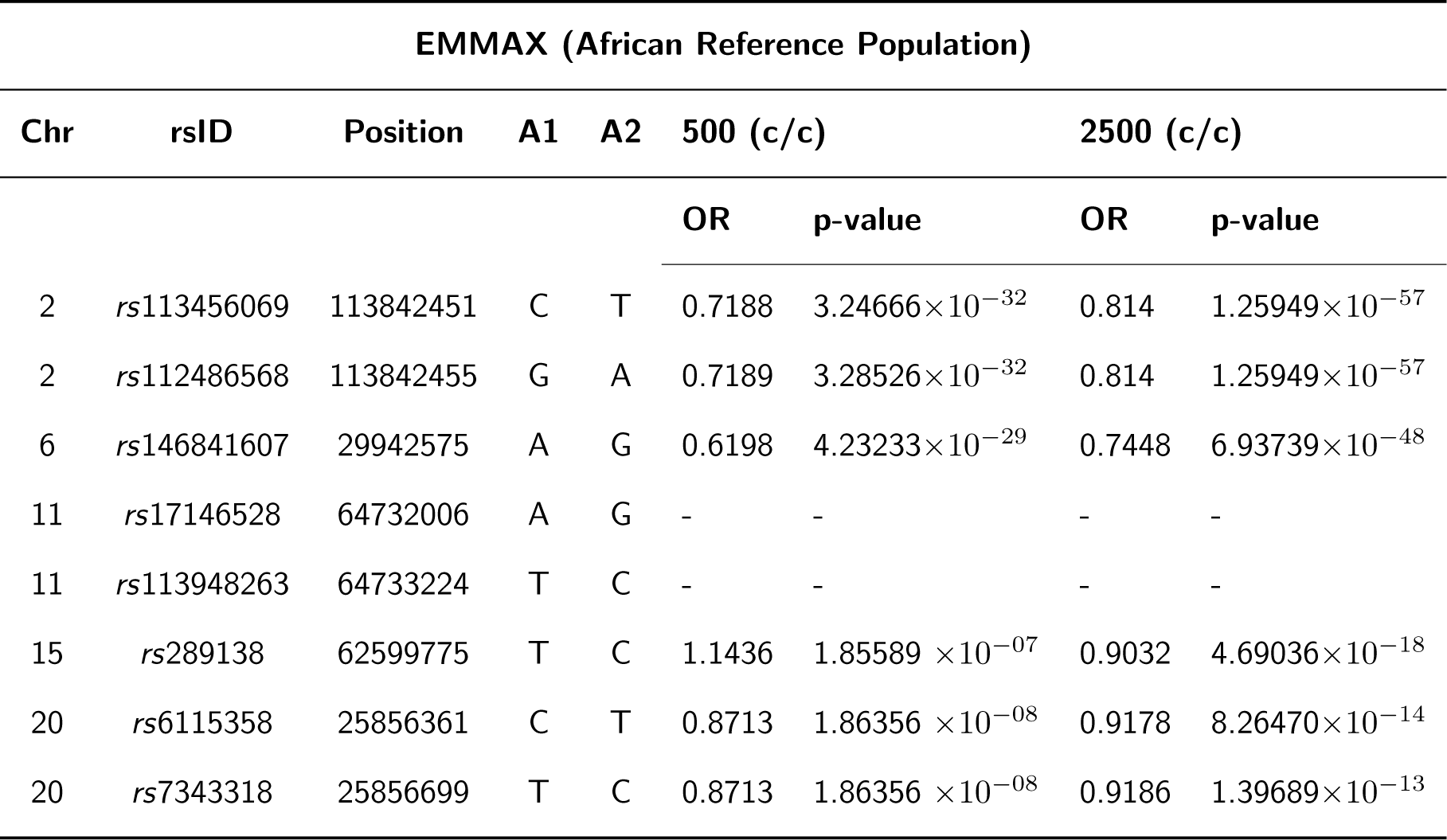
GWAS summary statistics from EMMAX of the simulated risk SNPs in the homogeneous simulation of 500 and 2500 samples from an African reference population of 221 samples. **x**(**c***/***c**) refers to the **x** number of cases/controls.

**Table 15:**
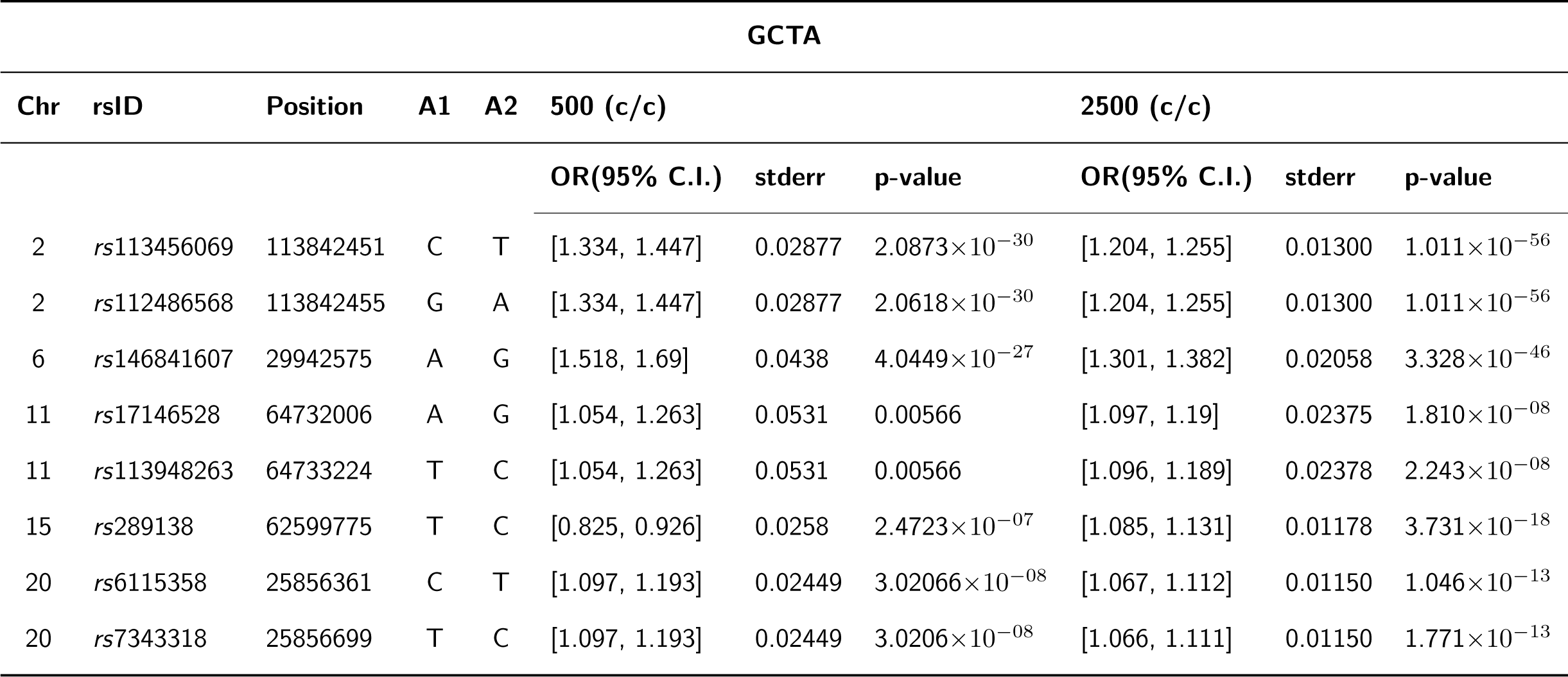
GWAS summary statistics from GCTA of the simulated risk SNPs in the homogeneous simulation of 500 and 2500 samples from an African reference population of 221 samples. **x**(**c***/***c**) refers to the **x** number of cases/controls.

**Table 16:**
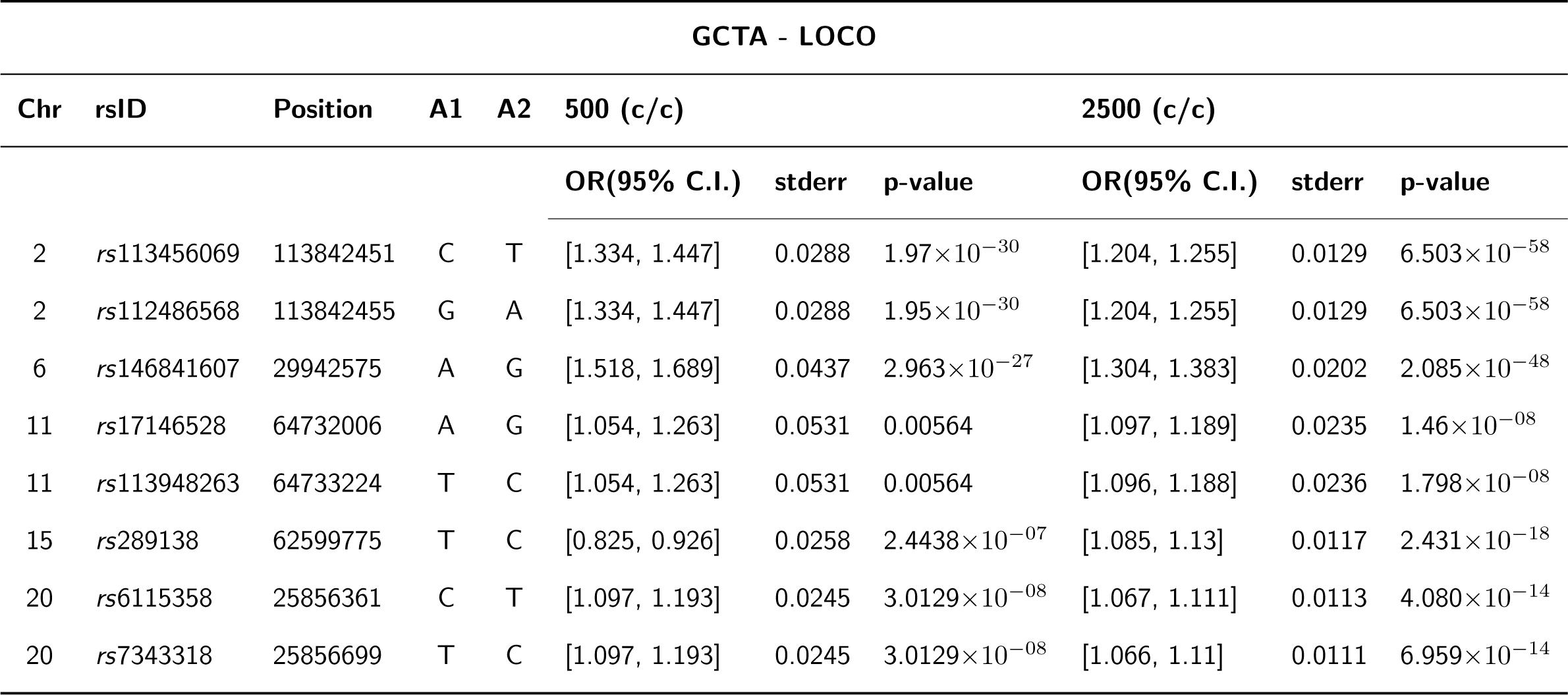
GWAS summary statistics from GCTA - LOCO of the simulated risk SNPs in the homogeneous simulation of 500 and 2500 samples from an African reference population of 221 samples. **x**(**c***/***c**) refers to the **x** number of cases/controls.

**Table 17:**
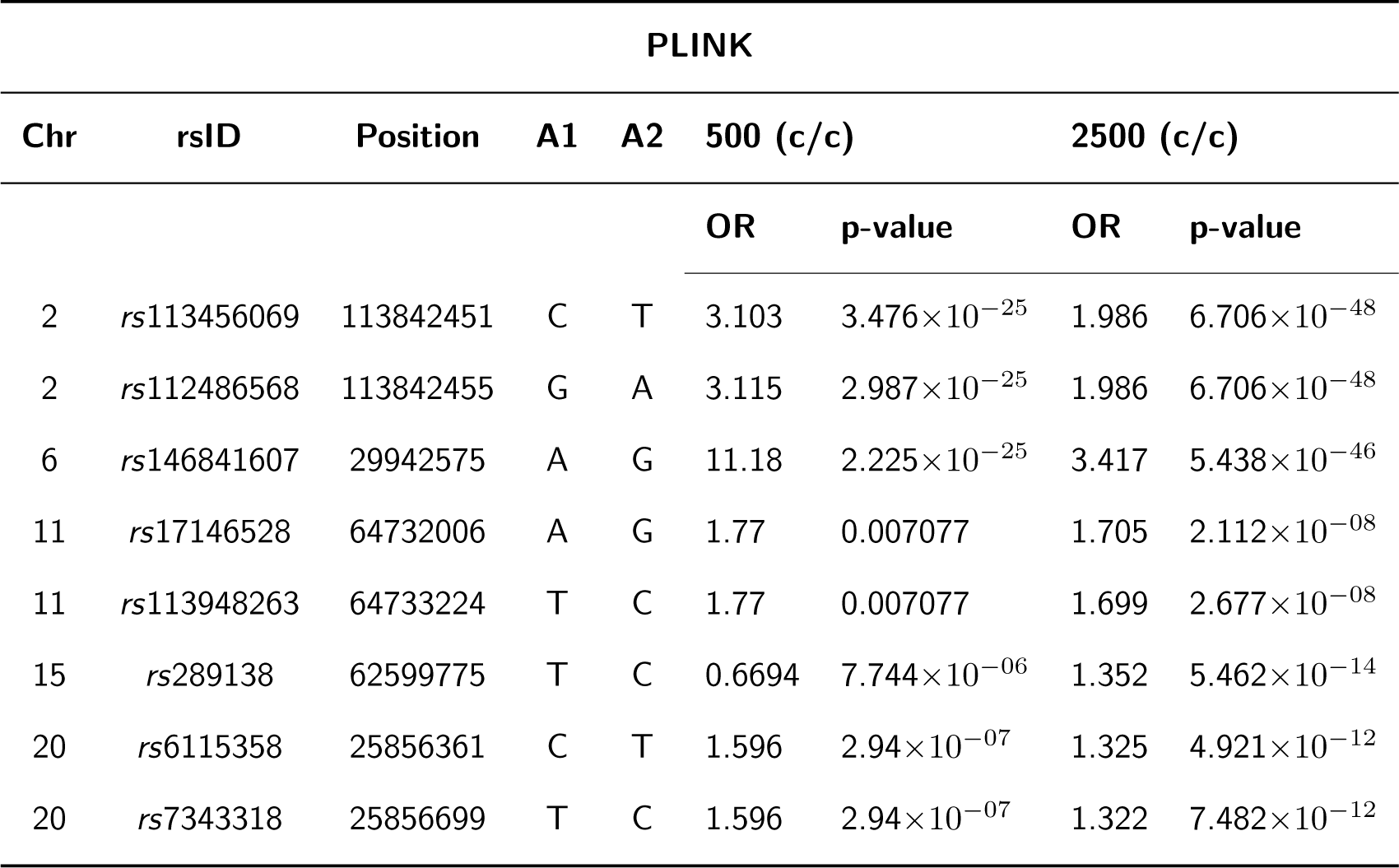
GWAS summary statistics from PLINK of the simulated risk SNPs in the homogeneous simulation of 500 and 2500 samples from an African reference population of 221 samples. **x**(**c***/***c**) refers to the **x** number of cases/controls.

**Table 18:**
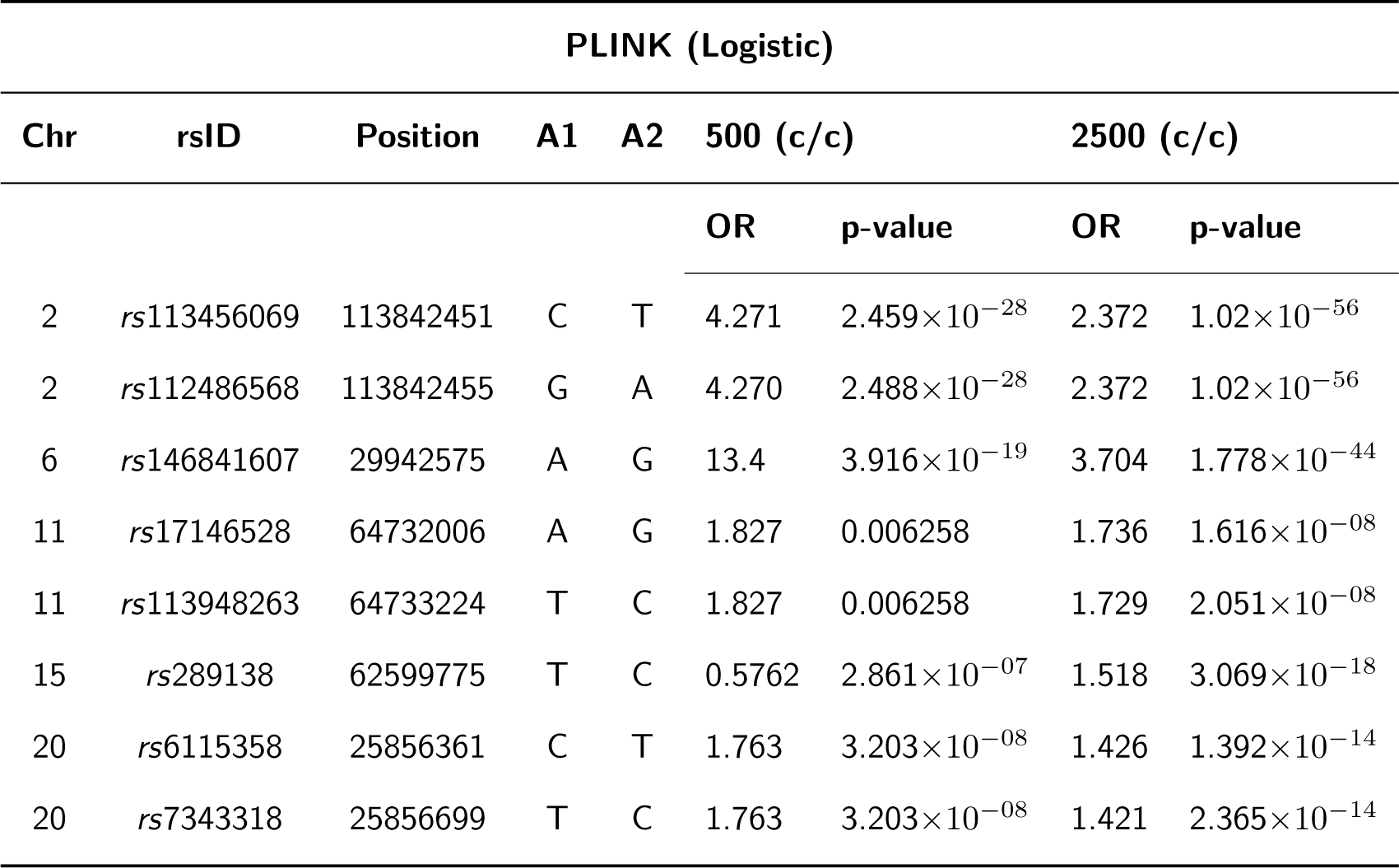
GWAS summary statistics from PLINK Logistic of the simulated risk SNPs in the homogeneous simulation of 500 and 2500 samples from an African reference population of 221 samples. **x**(**c***/***c**) refers to the **x** number of cases/controls.

**Table 19:**
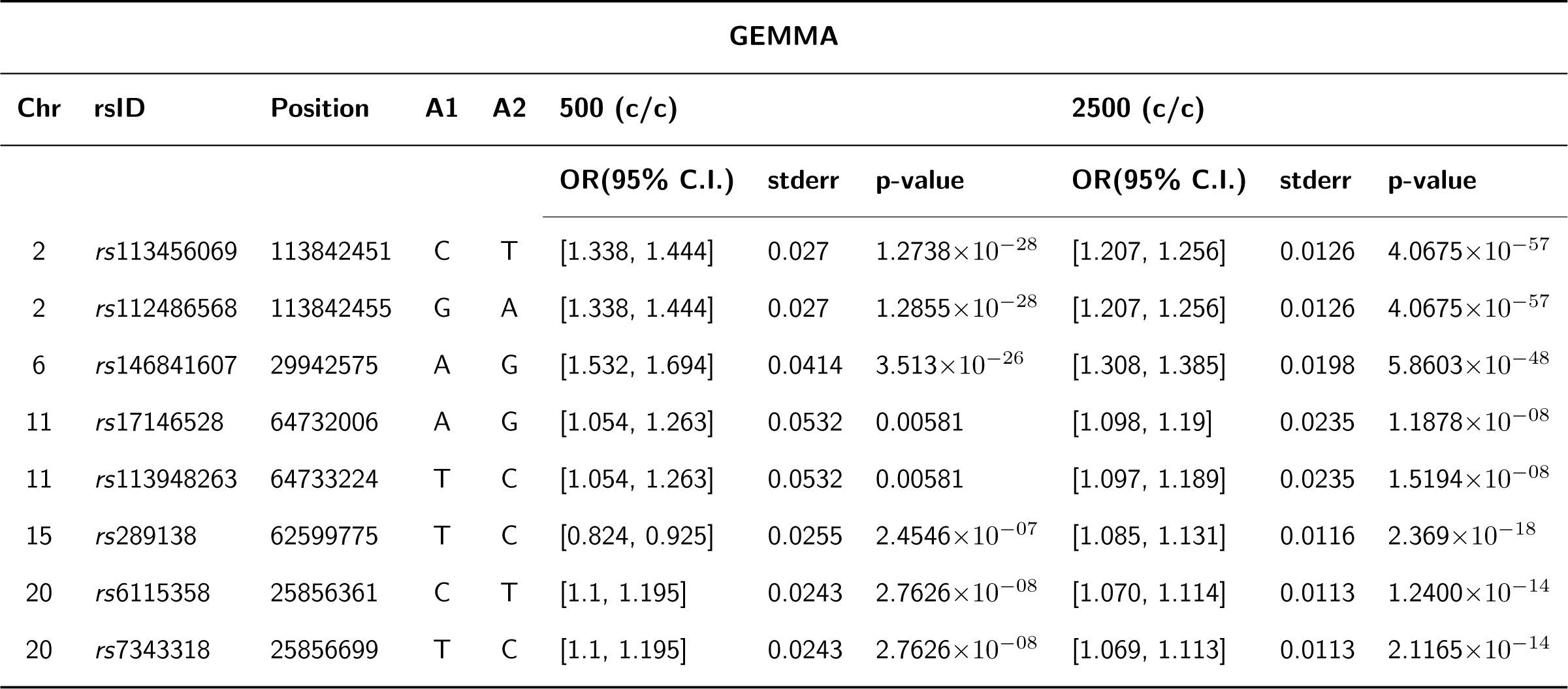
GWAS summary statistics from GEMMA of the simulated risk SNPs in the homogeneous simulation of 500 and 2500 samples from an African reference population of 221 samples. **x**(**c***/***c**) refers to the **x** number of cases/controls.

**Table 20:**
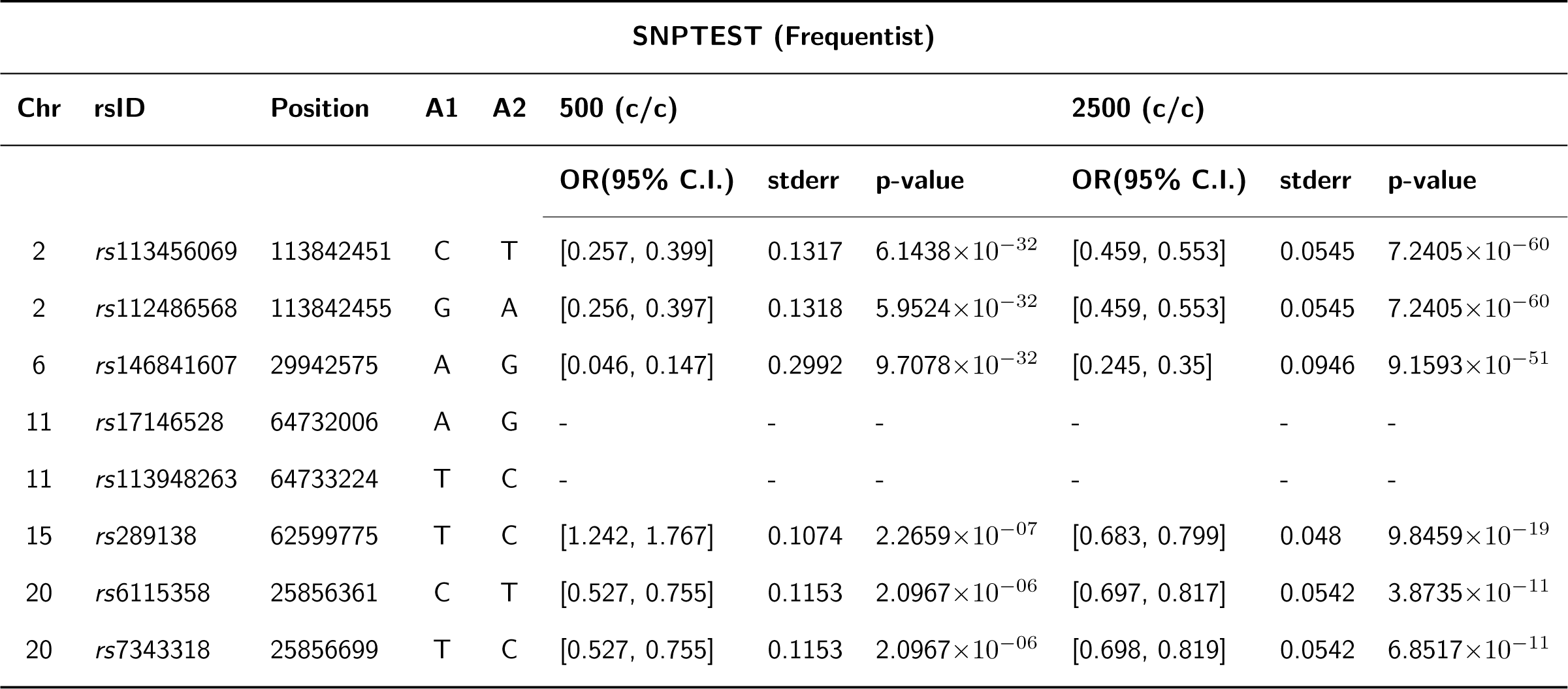
GWAS summary statistics from SNPTEST (Frequentist) of the simulated risk SNPs in the homogeneous simulation of 500 and 2500 samples from an African reference population of 221 samples. **x**(**c***/***c**) refers to the **x** number of cases/controls.

**Table 21:**
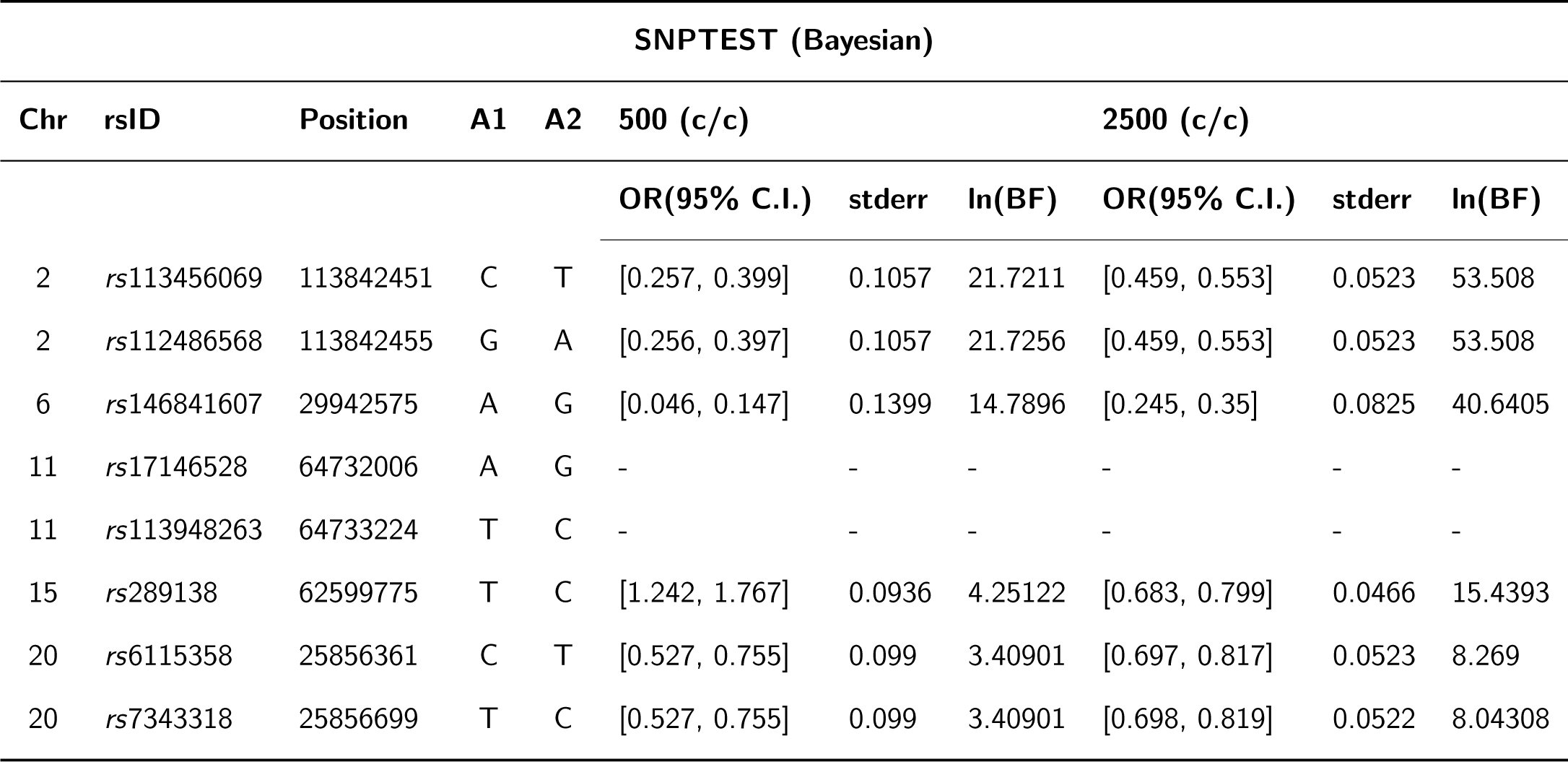
GWAS summary statistics from SNPTEST (Bayesian) of the simulated risk SNPs in the homogeneous simulation of 500 and 2500 samples from an African reference population of 221 samples. **x**(**c***/***c**) refers to the **x** number of cases/controls.

On increasing the sample size for the European population, we observed that all the tools were able to capture the simulated disease signals, and though EMMAX and SNPTEST excluded the risk SNP on chromosome 6 by internal quality control, SNPs in LD with these risk SNPs were captured for this population, and a significant signal was detected. However, in the African population, the signals at chromosomes 15 and 20 improved with increased sample sizes but were at a less significant threshold in comparison to the European population. We also note that at chromosome 11, where a weak signal was simulated for the African simulation, even with a larger sample size, the signals were still at a marginal significance thresholds with all 8 scoring statistics.

Our results thus suggested that in a homogeneous European population with small sample sizes, GEMMA, GCTA, and GCTA-LOCO were more robust in capturing most of the simulated risk variants at significant levels, with PLINK and PLINK-Logistic following suit. However, with large sample sizes, all the tools were effective in capturing the simulated risk at significance levels. We also noted that internal quality control checks implemented in EMMAX, SNPTEST-Frequentist, and SNPTEST-Bayesian that removes variants might remove risk variants, especially in analysis with small sample sizes, and thus missing out significant associations. However, our results revealed that with a small sample size, most tools were underpowered to detect some of the risk variants present at a significant level in the African GWAS, and even with an increased sample size, as observed on chromosome 11, some risk variants did struggle to reach the stringent GWAS threshold when the signal was weak.

Similar significant thresholds were observed for the risk SNPs simulated on chromosome 2, *rs*113456069 and *rs*112486568, in the European and African populations, respectively, and similarly on chromosome 20, *rs*6115358 and *rs*57343318, in the European and African populations, respectively. Of note is that only SNPs *rs*113456069 on chromosome 2 and *rs*6115358 on chromosome 20 were simulated as causal in the European population, while SNPs *rs*112486568 on chromosome 2 and *rs*7343318 on chromosome 20 were simulated in the African population. Though SNPs *rs*113456069 and *rs*112486568 on chromosome 2 were chosen to be in high LD in the European population, we observed that these SNPs were also in high LD in the African population. Similarly, SNPs *rs*6115358 and *rs*57343318 on chromosome 20 were also in high LD in the African datasets. We thus deduce that if strong risk signals exist in both European and African populations with high-powered studies, cross-population replication is possible using most of the tools assessed.

### Assessment of the Admixture Simulation GWAS Analysis

Figure 12 shows the Manhattan plots of the association tests of the seven disease scoring statistics assessed using the 3-way admixed population simulation, while **Tables 22** to **28** are the summary statistics of the risk SNPs simulated. We observed that the LMM-based tools EMMAX, GEMMA, and GCTA performed quite similarly in detecting the simulated risk variants and captured the risk variants on chromosomes 2 and 6 at a significant threshold. Though the three tools detected the risk variants simulated on chromosomes 11 and 15 at marginal significance thresholds, the SNPs in LD with the risk variant on chromosome 11 were detected as significant. GCTA-LOCO, an LMM-based approach, performed quite similarly to PLINK-Logistic, SNPTEST-Frequentist, and SNPTEST-Bayesian in capturing the risk variants on chromosomes 2, 6, and 11 as significant while capturing the signal on chromosome 15 at a marginal significance threshold. On chromosome 11, however, we note that the four approaches detected a second region that was not simulated with a risk variant and, thus, a false positive association that could be due to admixture. The four approaches also captured a significant signal on chromosome 12 that was not simulated as significant but detected at a marginal significance threshold by the other tools. We, therefore, noted that the LMM-based approaches EMMAX, GEMMA, and GCTA were more robust in capturing a wide range of population structures, which enabled them to control for any spurious associations. However, GCTA-LOCO, also an LMM-based approach, was ineffective in capturing the sample structures, and we hypothesize that the LOCO approach might have missed accounting for a significant amount of the sample structure in the analysis.

**Figure 12:**
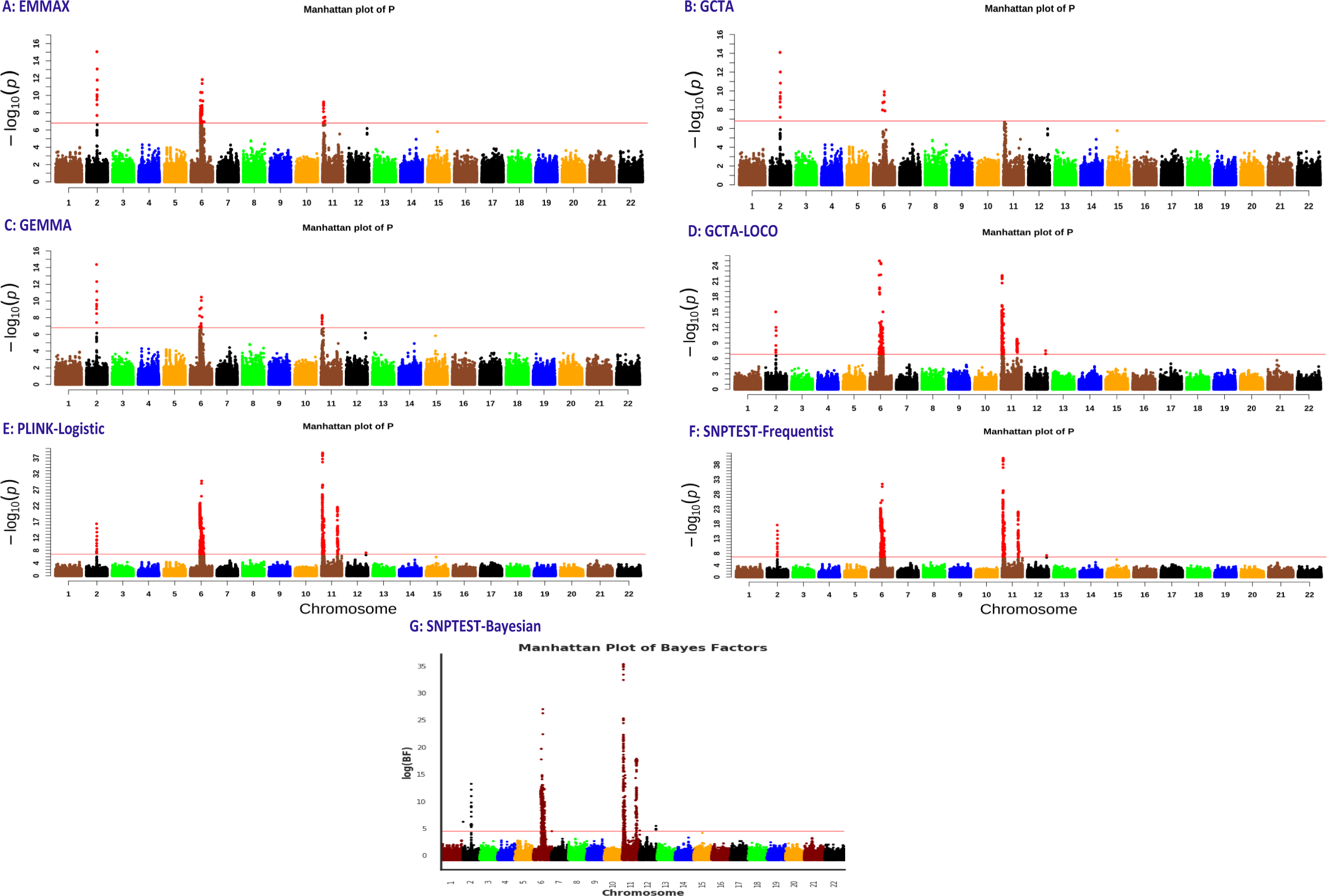
The Manhattan plots of GWAS of the simulated 3-way admixed population using EMMAX, GEMMA, PLINK, GCTA and SNPTEST tools. The red line represent the GWAS significance threshold line, and the significant SNPs are shown in red.

**Table 22:** GWAS statistics from EMMAX of the simulated risk SNPs in the 3-way admixture simulation for 2500 cases and 2500 controls.

| EMMAX |  |  |  |  |  |  |
| --- | --- | --- | --- | --- | --- | --- |
| Chr | rsID | Position | A1 | A2 | OR | p-value |
| 2 | rs76091761 | 119924776 | C | T | 0.8284 | $8.70874 \times 10^{-16}$ |
| 6 | rs79354975 | 78841154 | T | C | 0.9074 | $4.67783 \times 10^{-11}$ |
| 11 | rs73417185 | 4119431 | A | G | 0.9452 | $6.73071 \times 10^{-04}$ |
| 15 | rs2960806 | 60169020 | G | A | 1.0497 | $1.54807 \times 10^{-06}$ |

**Table 23:** GWAS statistics from GEMMA of the simulated risk SNPs in the 3-way admixture simulation for 2500 cases and 2500 controls.

| GEMMA |  |  |  |  |  |  |  |
| --- | --- | --- | --- | --- | --- | --- | --- |
| Chr | rsID | Position | A1 | A2 | OR(95% C.I.) | stderr | p-value |
| 2 | rs76091761 | 119924776 | C | T | [1.1627, 1.2533] | 0.02312 | $4.31377 \times 10^{-15}$ |
| 6 | rs79354975 | 78841154 | T | C | [1.0722, 1.1297] | 0.01467 | $6.39394 \times 10^{-10}$ |
| 11 | rs73417185 | 4119431 | A | G | [1.0232, 1.0879] | 0.0165 | $1.51937 \times 10^{-03}$ |
| 15 | rs2960806 | 60169020 | G | A | [0.9329, 0.9724] | 0.01009 | $1.46203 \times 10^{-06}$ |

**Table 24:** GWAS statistics from GCTA of the simulated risk SNPs in the 3-way admixture simulation for 2500 cases and 2500 controls.

| GCTA |  |  |  |  |  |  |  |
| --- | --- | --- | --- | --- | --- | --- | --- |
| Chr | rsID | Position | A1 | A2 | OR(95% C.I.) | stderr | p-value |
| 2 | rs76091761 | 119924776 | C | T | [1.1573, 1.2511] | 0.02392 | $7.99158 \times 10^{-15}$ |
| 6 | rs79354975 | 78841154 | T | C | [1.0651, 1.1236] | 0.01492 | $1.52098 \times 10^{-09}$ |
| 11 | rs73417185 | 4119431 | A | G | [1.0182, 1.082] | 0.01628 | $2.68599 \times 10^{-03}$ |
| 15 | rs2960806 | 60169020 | G | A | [0.9329, 0.9726] | 0.01012 | $1.69894 \times 10^{-06}$ |

**Table 25:** GWAS statistics from GCTA-LOCO of the simulated risk SNPs in the 3-way admixture simulation for 2500 cases and 2500 controls.

| GCTA-LOCO |  |  |  |  |  |  |  |
| --- | --- | --- | --- | --- | --- | --- | --- |
| Chr | rsID | Position | A1 | A2 | OR(95% C.I.) | stderr | p-value |
| 2 | rs76091761 | 119924776 | C | T | [1.1378, 1.2175] | 0.02034 | $8.93609 \times 10^{-16}$ |
| 6 | rs79354975 | 78841154 | T | C | [1.0972, 1.142] | 0.01144 | $5.40953 \times 10^{-23}$ |
| 11 | rs73417185 | 4119431 | A | G | [1.0656, 1.111] | 0.01158 | $2.66512 \times 10^{-13}$ |
| 15 | rs2960806 | 60169020 | G | A | [0.9488, 0.9844] | 0.00908 | 0.00018 |

**Table 26:**
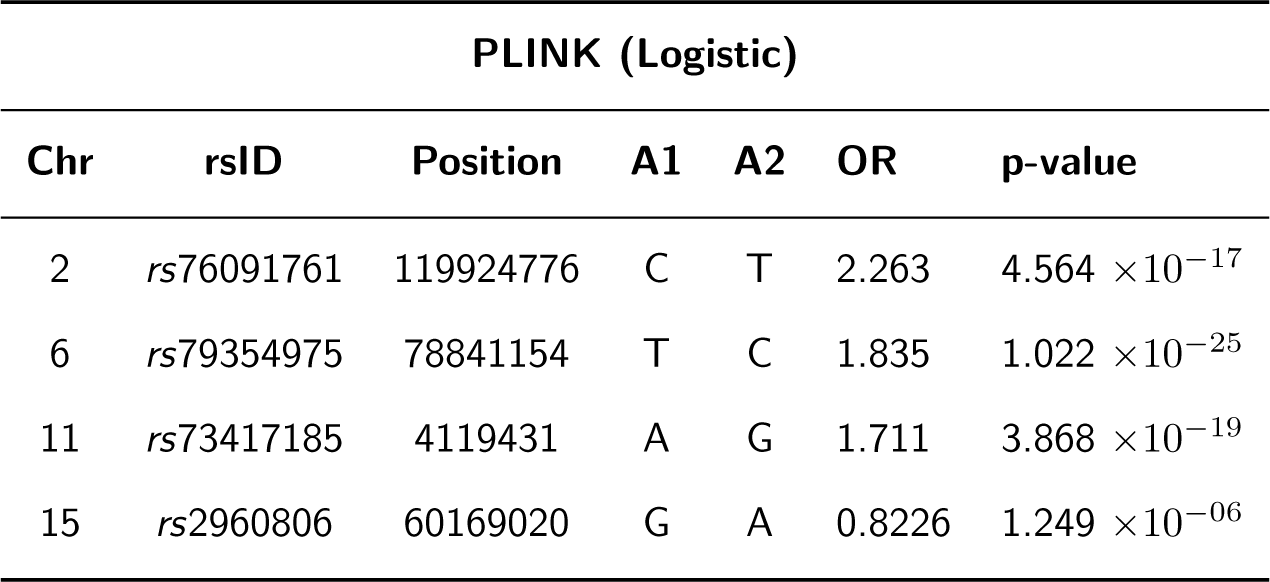
GWAS statistics from PLINK (Logistic) of the simulated risk SNPs in the 3-way admixture simulation for 2500 cases and 2500 controls.

**Table 27:** GWAS statistics from SNPTEST of the simulated risk SNPs in the 3-way admixture simulation for 2500 cases and 2500 controls.

| SNPTTEST - Frequentist |  |  |  |  |  |  |  |
| --- | --- | --- | --- | --- | --- | --- | --- |
| Chr | rsID | Position | A1 | A2 | OR(95% C.I.) | stderr | p-value |
| 2 | rs76091761 | 119924776 | C | T | [0.2517, 0.6329] | 0.09724 | $2.49893 \times 10^{-18}$ |
| 6 | rs79354975 | 78841154 | T | C | [0.43, 0.6571] | 0.05793 | $1.0629 \times 10^{-26}$ |
| 11 | rs73417185 | 4119431 | A | G | [0.4674, 0.7028] | 0.06005 | $1.47774 \times 10^{-19}$ |
| 15 | rs2960806 | 60169020 | G | A | [1.1367, 1.2946] | 0.04028 | $1.18143 \times 10^{-06}$ |

**Table 28:** GWAS statistics from SNPTEST of the simulated risk SNPs in the 3-way admixture simulation for 2500 cases and 2500 controls.

| SNPTEST - Bayesian |  |  |  |  |  |  |  |
| --- | --- | --- | --- | --- | --- | --- | --- |
| Chr | rsID | Position | A1 | A2 | OR(95% C.I.) | stderr | log(BF) |
| 2 | rs76091761 | 119924776 | C | T | [0.3474, 0.683] | 0.08562 | 13.2702 |
| 6 | rs79354975 | 78841154 | T | C | [0.4611, 0.6782] | 0.05539 | 22.4432 |
| 11 | rs73417185 | 4119431 | A | G | [0.4992, 0.7238] | 0.0573 | 15.815 |
| 15 | rs2960806 | 60169020 | G | A | [1.129, 1.2838] | 0.03947 | 4.22825 |

In the assessment of the tools using a 5-way admixed population, we obtained the Manhattan plots on Figures 13 and 14 and the summary statistics of the risk SNPs simulated on **Tables 29** to **35** on page 42. We observed that for the small sample size of 500 cases and 500 controls, all the tools could capture the simulated risk variants on chromosomes 2, 6, and 20. However, none of the tools captured the risk variants on chromosomes 11 and 15 at a significant level. With a large sample size, we observed that all the tools could capture one of the risk variants on chromosome 11, but the signal at chromosome 15 could still not reach the significant threshold.

**Figure 13:**
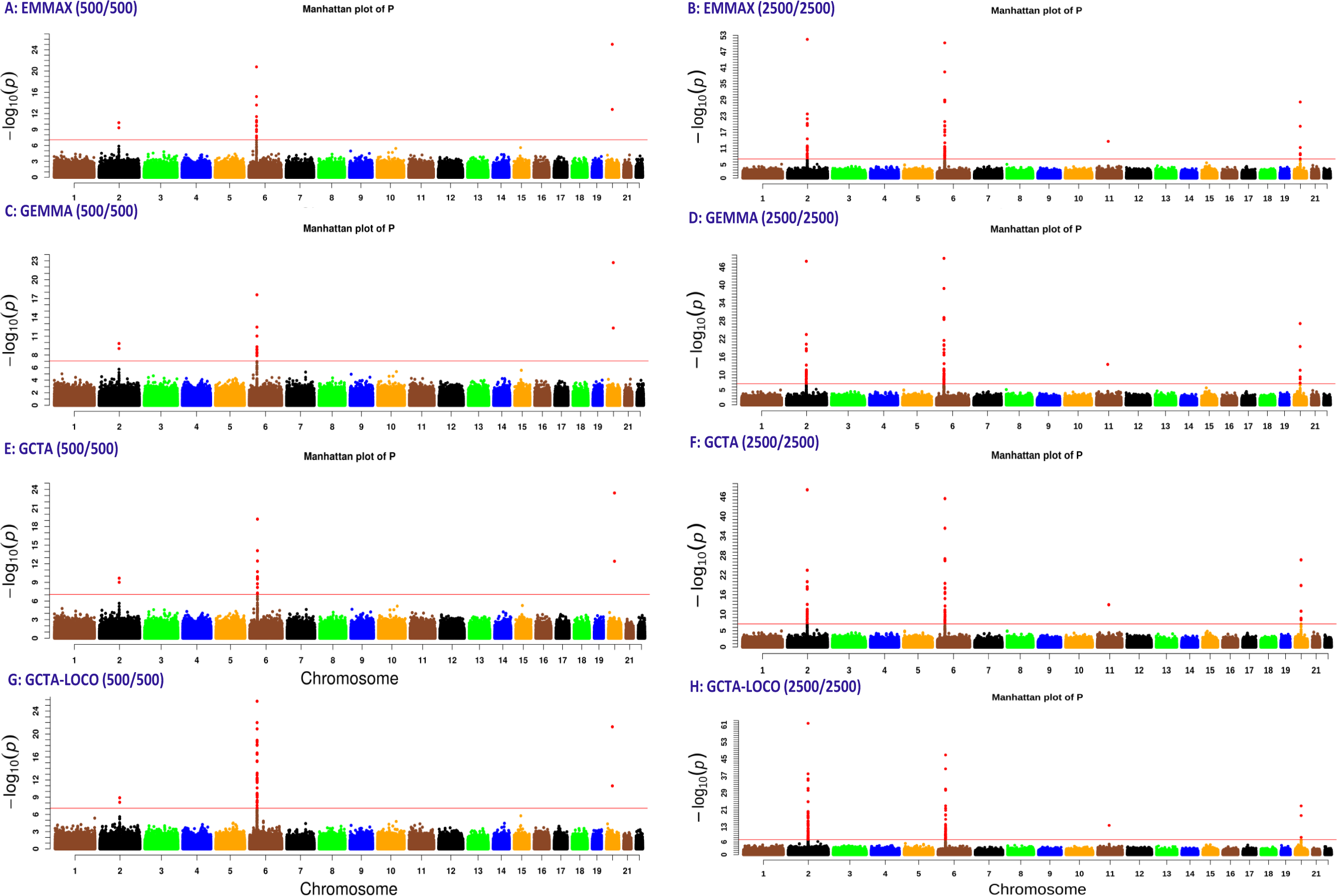
The Manhattan plots of GWAS of the simulated 5-way admixed population using EMMAX, GEMMA, and GCTA. Column A-C-E-G correspond to the 500 cases and 500 controls simulation, while column B-D-F-H correspond to 2500 cases and 2500 controls simulation. The significance threshold line and the significant SNPs are shown in red.

**Figure 14:**
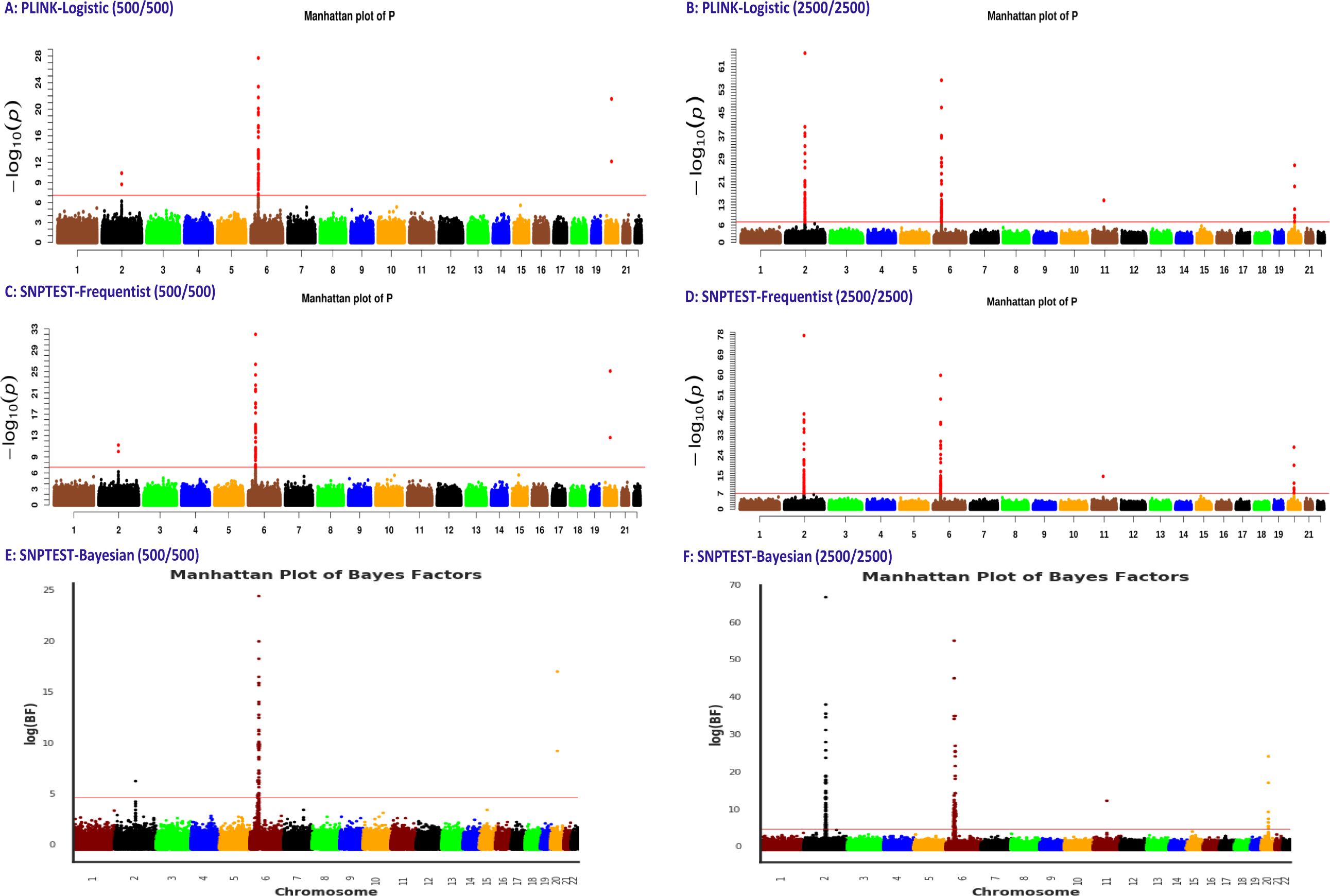
The Manhattan plots of GWAS of the simulated 5-way admixed population using PLINK and SNPTEST. Column A-C-E correspond to the 500 cases and 500 controls simulation, while column B-D-F correspond to 2500 cases and 2500 controls simulation. The significance threshold line and the significant SNPs are shown in red.

**Table 29:**
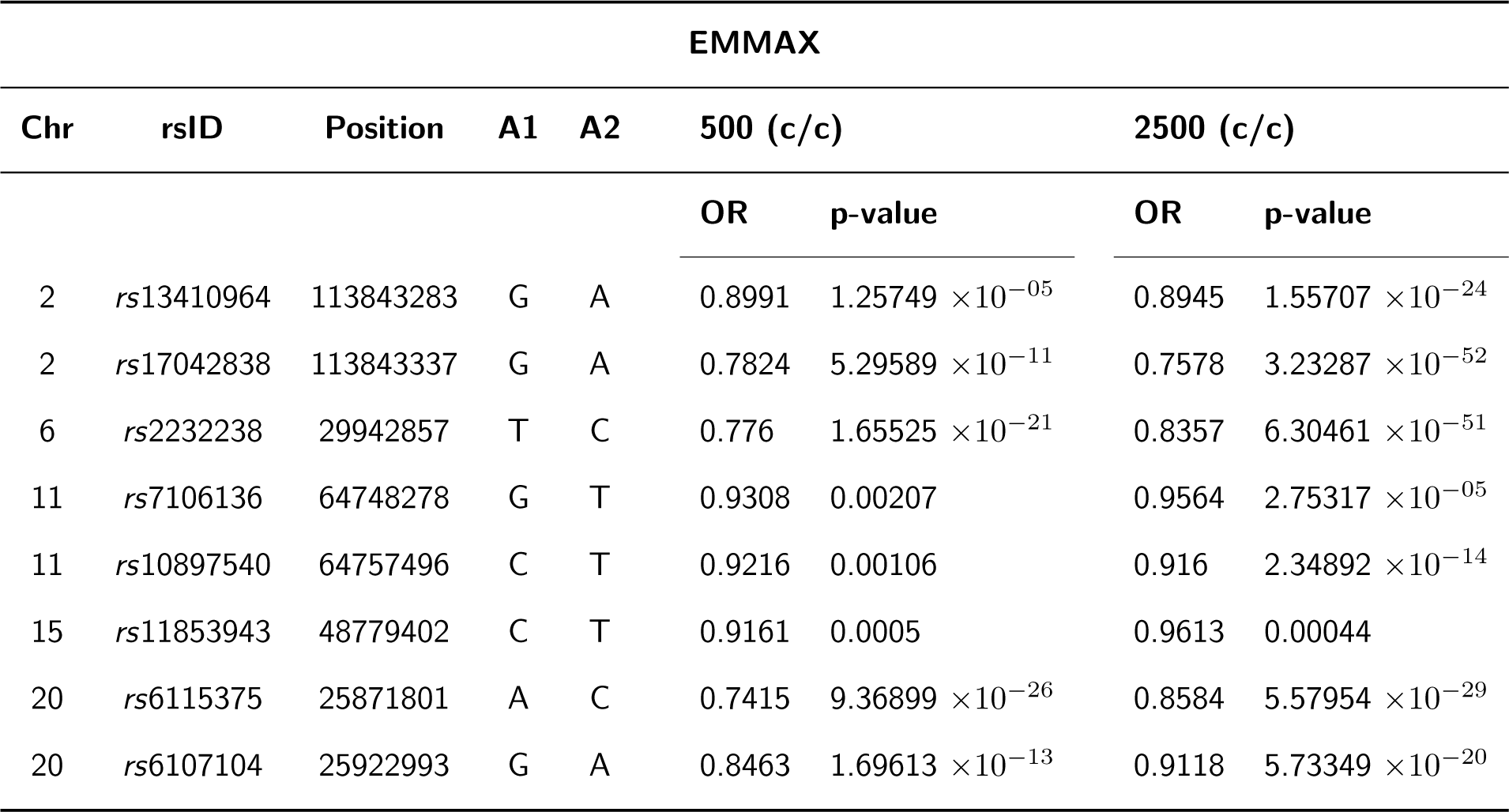
GWAS statistics from EMMAX of the simulated risk SNPs in the 5-way admixture simulation for the 500 cases and 500 controls (500 (c/c)) and 2500 cases and the 2500 controls (2500 (c/c)) sample sizes.

**Table 30:**
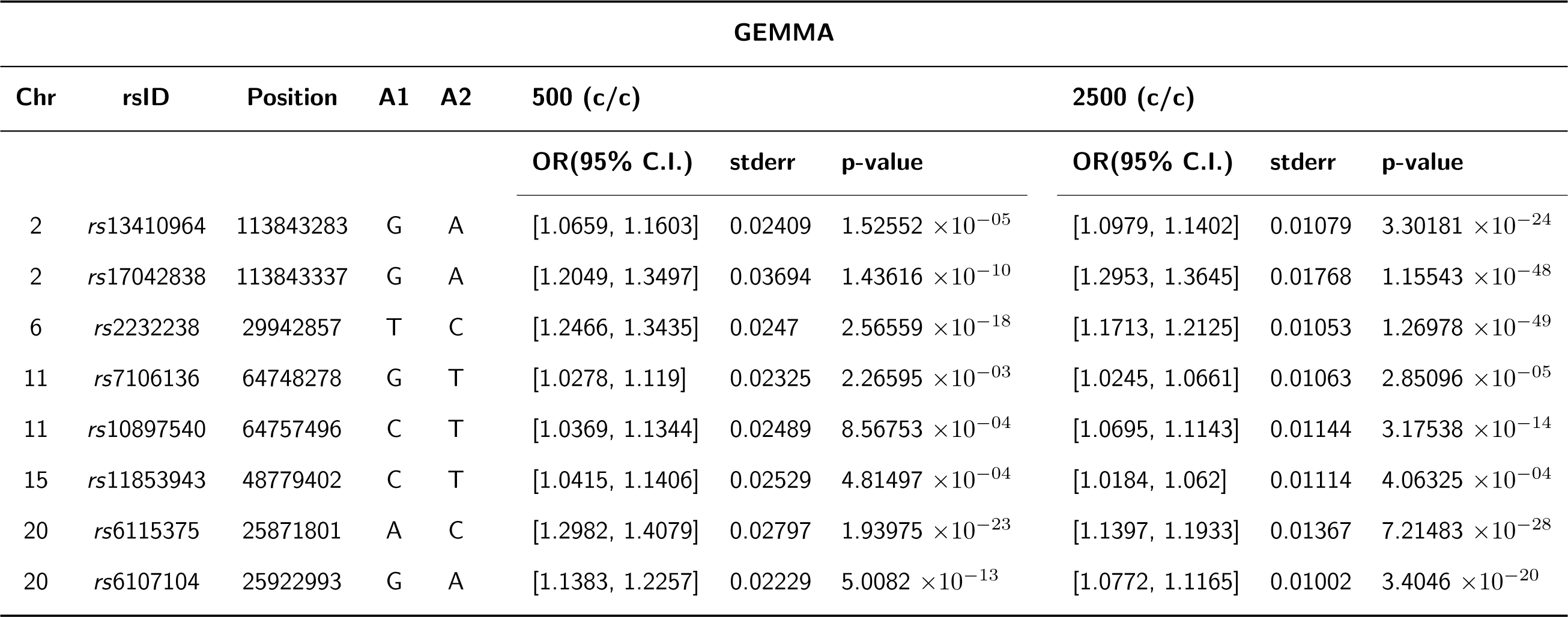
GWAS statistics from GEMMA of the simulated risk SNPs in the 5-way admixture simulation for the 500 cases and 500 controls (500 (c/c)) and 2500 cases and the 2500 controls (2500 (c/c)) sample sizes.

**Table 31:**
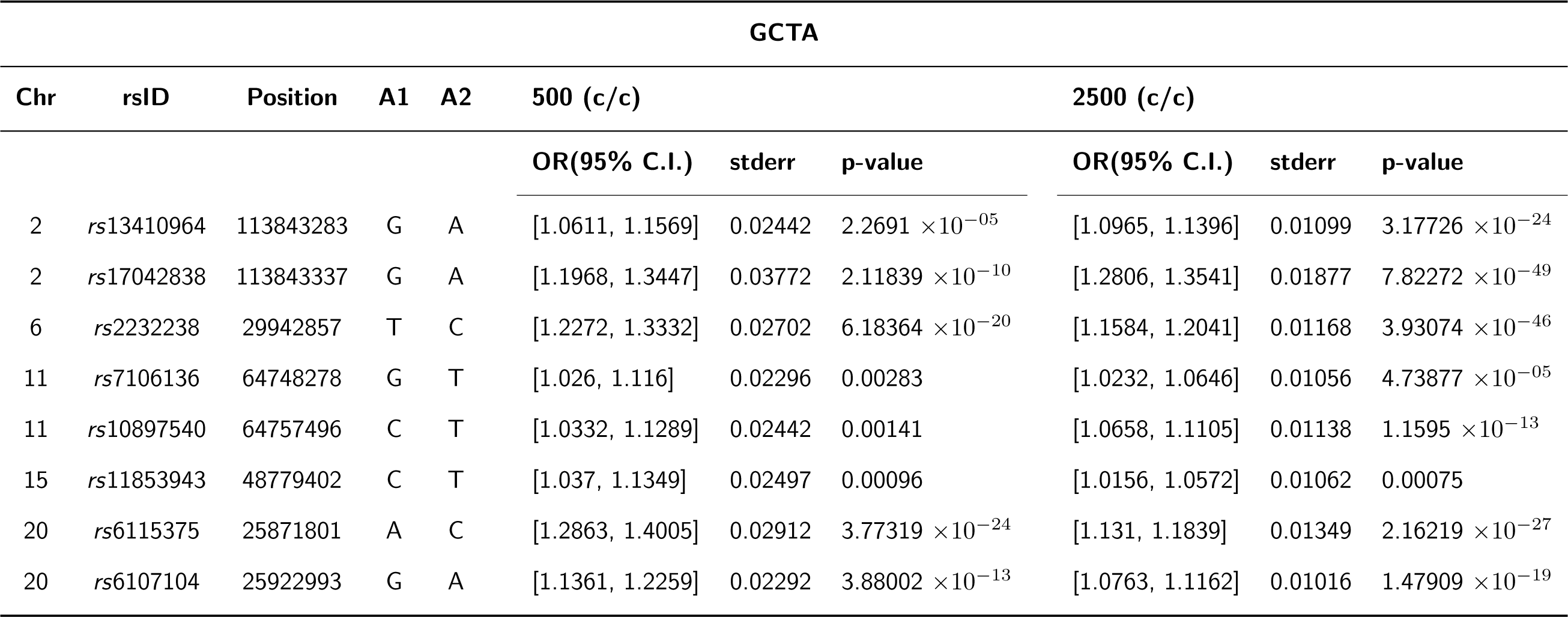
GWAS statistics from GCTA of the simulated risk SNPs in the 5-way admixture simulation for the 500 cases and 500 controls (500 (c/c)) and 2500 cases and the 2500 controls (2500 (c/c)) sample sizes.

**Table 32:**
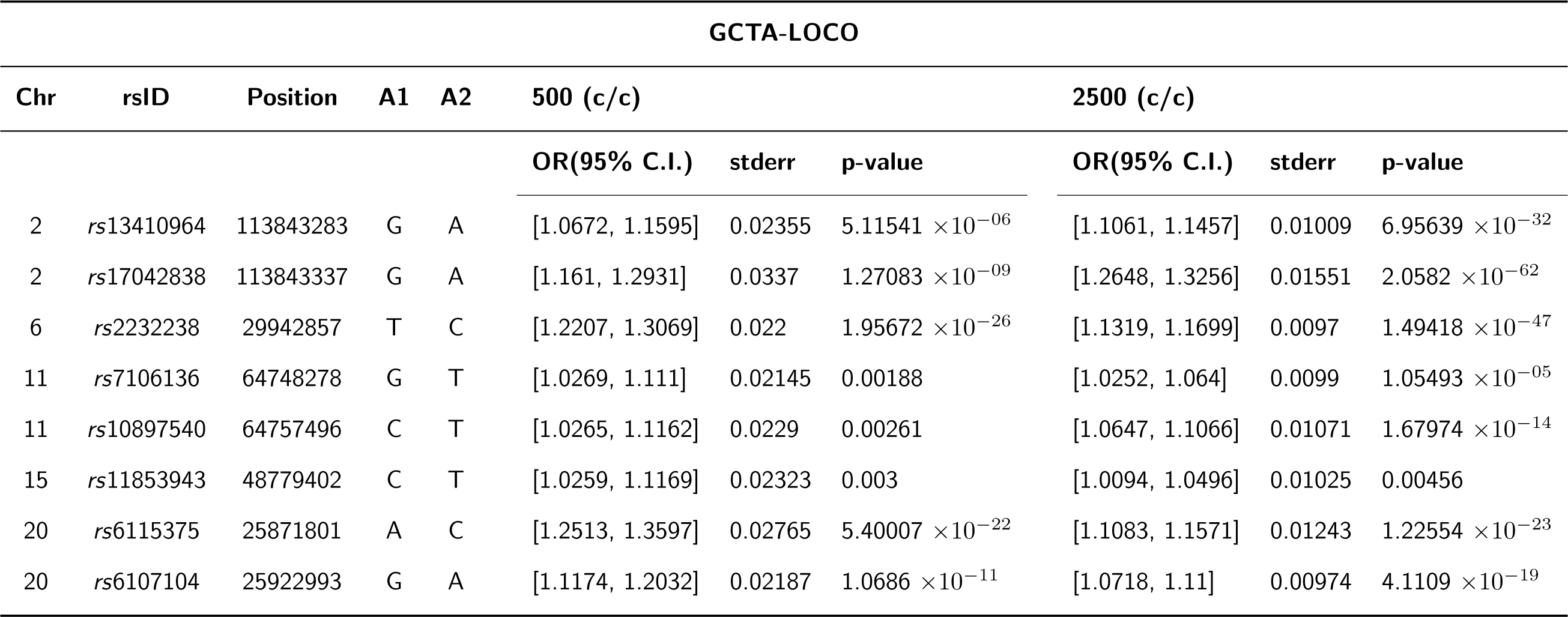
GWAS statistics from GCTA-LOCO of the simulated risk SNPs in the 5-way admixture simulation for the 500 cases and 500 controls (500 (c/c)) and 2500 cases and the 2500 controls (2500 (c/c)) sample sizes.

**Table 33:**
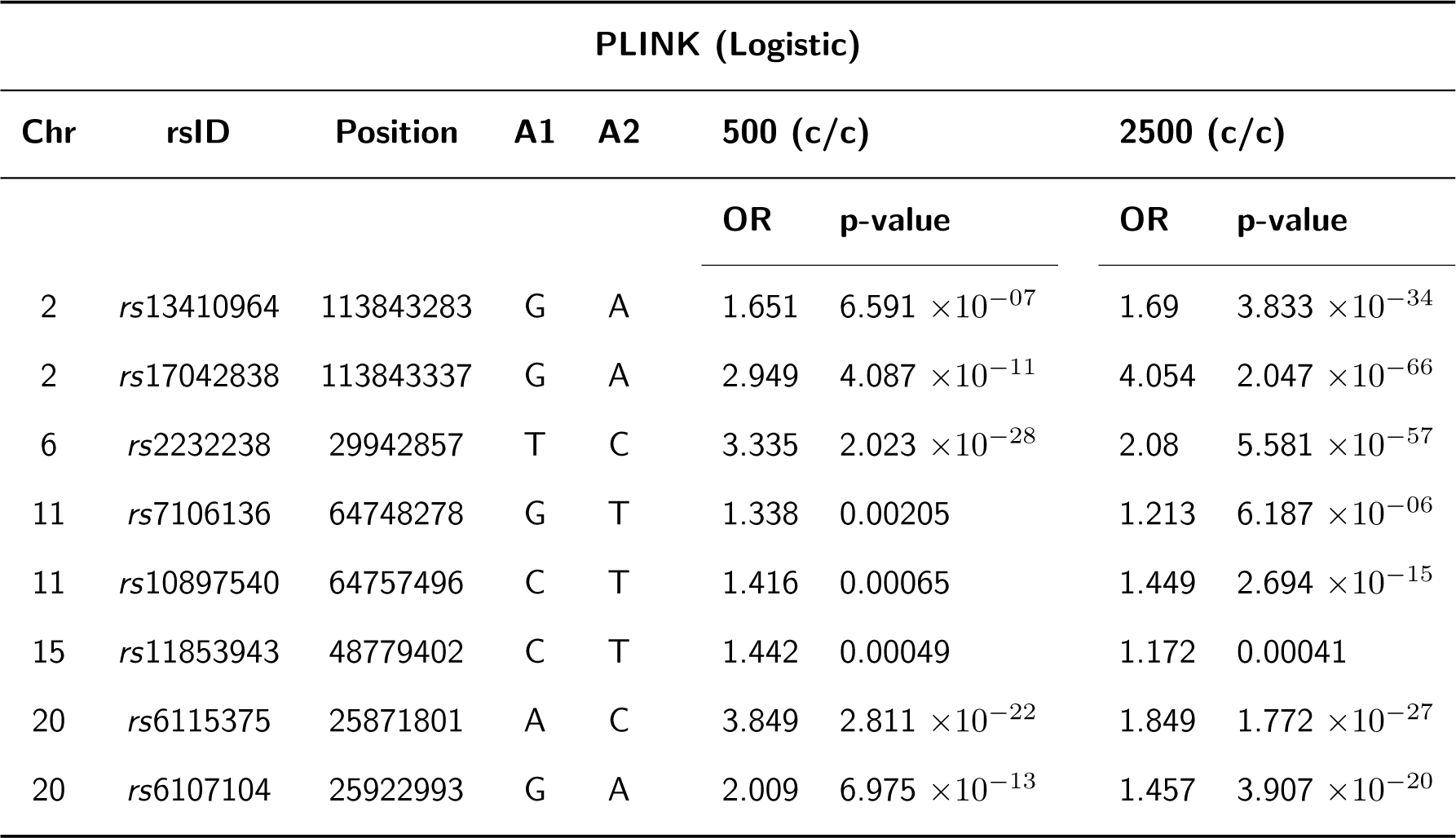
GWAS statistics from PLINK (Logistic) of the simulated risk SNPs in the 5-way admixture simulation for the 500 cases and 500 controls (500 (c/c)) and 2500 cases and the 2500 controls (2500 (c/c)) sample sizes.

**Table 34:**
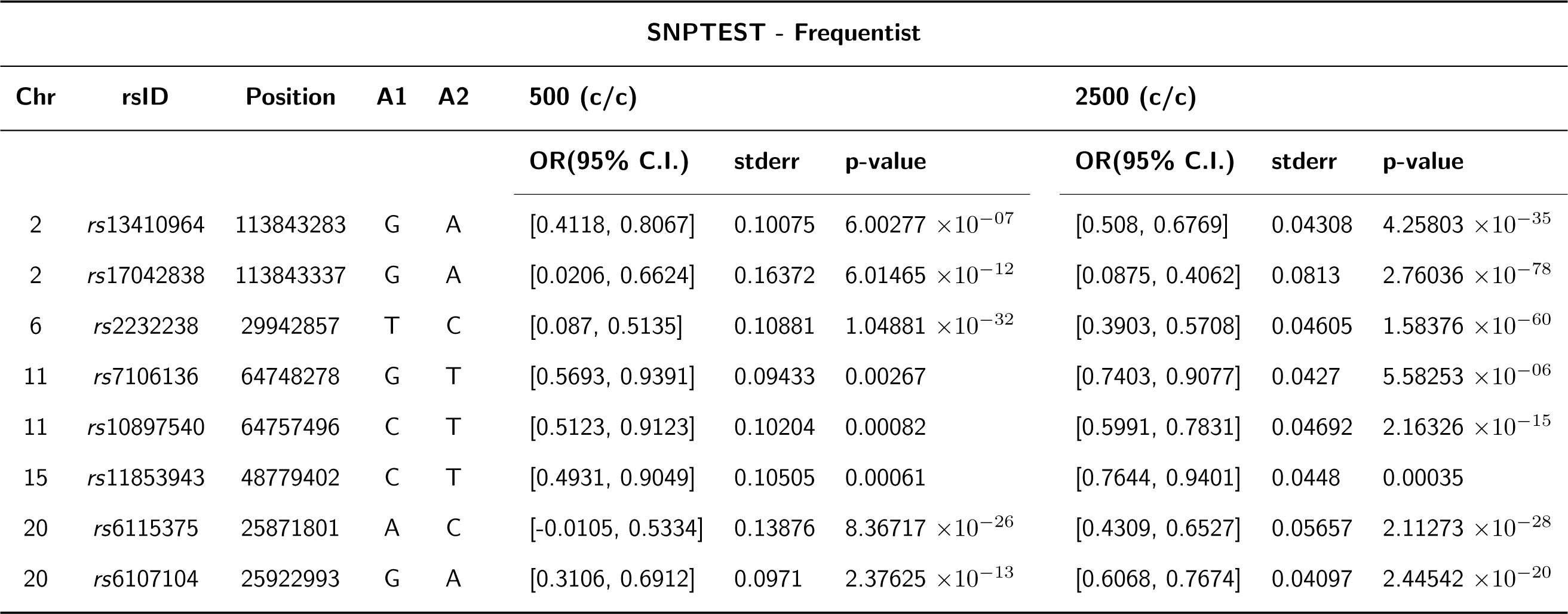
GWAS statistics from SNPTEST of the simulated risk SNPs in the 5-way admixture simulation for the 500 cases and 500 controls (500 (c/c)) and 2500 cases and the 2500 controls (2500 (c/c)) sample sizes.

**Table 35:** GWAS statistics from SNPTEST of the simulated risk SNPs in the 5-way admixture simulation for the 500 cases and 500 controls (500 (c/c)) and 2500 cases and the 2500 controls (2500 (c/c)) sample sizes.

| SNPTEST - Bayesian |  |  |  |  |  |  |  |  |  |  |
| --- | --- | --- | --- | --- | --- | --- | --- | --- | --- | --- |
| Chr | rsID | Position | A1 | A2 | 500 (c/c) |  |  | 2500 (c/c) |  |  |
|  |  |  |  |  | OR(95% C.I.) | stderr | log(BF) | OR(95% C.I.) | stderr | log(BF) |
| 2 | rs13410964 | 113843283 | G | A | [0.4981, 0.8478] | 0.08923 | 4.01393 | [0.524, 0.6886] | 0.042 | 31.1146 |
| 2 | rs17042838 | 113843337 | G | A | [0.2777, 0.7571] | 0.12231 | 6.27021 | [0.1564, 0.4418] | 0.0728 | 66.6651 |
| 6 | rs2232238 | 29942857 | T | C | [0.2111, 0.5701] | 0.09159 | 24.4739 | [0.411, 0.586] | 0.04463 | 55.13 |
| 11 | rs7106136 | 64748278 | G | T | [0.627, 0.9606] | 0.08512 | 1.24261 | [0.7492, 0.9128] | 0.04175 | 3.60867 |
| 11 | rs10897540 | 64757496 | C | T | [0.5863, 0.9413] | 0.09056 | 1.60045 | [0.6151, 0.794] | 0.04562 | 12.3313 |
| 15 | rs11853943 | 48779402 | C | T | [0.5734, 0.9367] | 0.09267 | 1.68192 | [0.7731, 0.9444] | 0.0437 | 1.98332 |
| 20 | rs6115375 | 25871801 | A | C | [0.1833, 0.6061] | 0.10785 | 17.051 | [0.4607, 0.673] | 0.05416 | 24.1036 |
| 20 | rs6107104 | 25922993 | G | A | [0.4017, 0.7389] | 0.08601 | 9.23123 | [0.619, 0.7761] | 0.04008 | 17.1318 |

We thus noted that when the genotype risk was strong, irrespective of the presence and strength of the ancestry association, all the tools were also able to detect the risk variant at a significant level, as observed on chromosomes 2 and 6 in the 3-way simulation and on chromosomes 2, 6, and 20 in the 5-way simulation analysis. This was true for most tools, even with the smaller sample size in the 5-way simulation analysis. However, when the genotype risk was weak and the ancestry risk present was weak or strong, most of the tools were limited in their ability to detect the simulated risk variant at a significant level, as observed on chromosome 15 in the 3-way simulation and on chromosomes 11 and 15 in the 5-way simulation. Though GCTA-LOCO, PLINK-Logistic, SNPTEST-Frequentist, and SNPTEST-Bayesian were able to detect the risk SNP simulated on chromosome 11 in the 3-way admixed simulation at significance thresholds, they were limited in capturing the admixture-LD on this chromosome and resulted in spurious association signals, which GEMMA, EMMAX, and GCTA were successful in controlling for; however, they detected this risk variant at marginal significance thresholds. By increasing the sample size, one simulated risk SNP on chromosome 11 in the 5-way admixed population association was also detected as significant by all tools. The simulated ancestry risk on this chromosome was weak, which implied that the increase in power to detect the risk variant was highly likely due to the increase in sample size and not associated with ancestry risk.

## Discussion

In this study, we implemented FractalSIM and simulated European, African, and admixed populations to evaluate five of the commonly used GWAS tools on their performance in GWAS of diverse populations. Our results suggested that LMM-based tools were more robust in capturing risk variants present in the European population with smaller samples, but with increased samples, all the tools performed similarly. In the African population, all the tools were limited in their ability to capture risk variants present in small sample sizes. Though increasing the sample size did improve the power to capture the risk variants, when the signal was weak, some risk variants still struggled to reach the significant levels set in GWAS. The standard significance threshold for GWAS, 5.0 *×* 10*^−^*^08^, has been set using the European population. Given the increased independent testing in African population GWAS analysis due to generally higher number of SNPs and short LD blocks, it has been suggested and shown that a stricter significance threshold should be considered (Pulit et al., 2017). Taking this into account, it raises the question of whether the risk signals observed at the near marginal significance thresholds in our African population study with increased sample sizes would still be significant with more stringent thresholds. This, therefore, emphasizes the dire need for increased sampling in African populations if African GWAS is to catch up with European GWAS, given that small sample sizes still plague African GWAS.

Our homogeneous GWAS analysis also showed that, using most of the tools assessed, there is a possibility of cross-population replication in the presence of strong risk signals in both European and African populations when the studies are high-powered. However, caution should be exercised while using EMMAX, SNPTEST-Frequentist, and SNPTEST-Bayesian approaches, as we noted that internal quality controls in the tools could eliminate risk variants in the analysis.

In the admixture context, we also observed that the LMM-based models, except for GCTA-LOCO, performed better in controlling for spurious associations. However, they were limited in detecting the simulated risk variant when the genotype risk was very weak, irrespective of whether the ancestry risk was very high or moderate at the genomic region containing the risk SNP. Though increasing the sample size improved the power to capture some risk variants using all the tools, similar to the African population, some risk variants still struggled to reach the significant threshold. Therefore, as GWAS extends to diverse populations, it should be noted that, though increasing sampling in admixed populations may improve the power to detect some variants in the population using the tools assessed, association methods that can leverage ancestry risk in multi-way admixed populations, as has been illustrated in GWAS of 2-way admixed populations, will play a key role in improving GWAS power in these populations.

Consistent with other recent studies, the lesson learnt from the various GWAS in our study is that one should consider (i) applying population-specific GWAS pipelines and significance thresholds; (ii) choosing appropriate GWAS tools among the existing tools or possibly running multiple GWAS tools to allow a genome-wide level of significance to have consensus across many tools; (iii) the direction of effect size in each study in meta-analysis with diverse populations to replicate European GWAS and (iv) reporting population specific minor allele frequency, effect size, standard error of the effect size and LD of the associated variants in diverse populations to enable improved interpretation of the results.

The high genetic diversity of African and other diverse populations may enable the detection of many novel variants that are yet to be described in current public databases, such as the GWAS catalog (Buniello et al., 2019) or PhenGenI (Pasha and Scaria, 2013). It is thus important to develop new or adapted pipelines for diverse genetic data or to benchmark existing bioinformatics pipeline tools using diverse populations to account for diverse genetic and environmental characteristics that could differently shape phenotypic variation.

Numerous studies have leveraged local-specific ancestry tracts in variant-level association analyses for African Americans (Kim et al., 2022), Latinos (Torgerson et al., 2012), South African Coloured (Chimusa et al., 2014) and Hispanic cohorts (Kizil et al., 2022), demonstrating added value beyond standard association testing. Admixture association critically relies on accurate local ancestry inference (LAI), which requires well-specified founding population reference samples (Shriner, 2017). Though combining admixture mapping and SNP association testing has been shown to improve power in GWAS (Shriner et al., 2011; Salter-Townshend and Myers, 2019), this approach is rarely adopted because of the multi-stage process required and the challenge of application to complex admixed samples (Thornton and Bermejo, 2014), while most joint approaches to date are tailored to 2-way admixed populations (Tang et al., 2010; Pasaniuc et al., 2011; Shriner et al., 2011; Atkinson et al., 2021). Therefore, there is a critical need to (1) improve LAI accuracy (Geza et al., 2019), (2) build integrative software for running multi-way admixture deconvolution analysis (Geza et al., 2020), (3) design user-friendly, integrative joint association methods that generate comprehensive association statistics, and (4) optimize the power of association testing in multi-way admixed data (Duncan et al., 2019; Coram et al., 2017; Marnetto et al., 2020).

## Statements and Declarations

## Data Availability

The data used in this study is publicly available in the 1000 Genomes catalog. Simulated data is also available by request from the authors.

https://www.internationalgenome.org/

## Acknowledgments

We acknowledge the High Performance Computing resources at the University of Cape Town and the National Integrated CyberInfrastructure System (NICIS) for providing access to their Centre for High Performance Computing resources that were used to run all the analysis in this research work.

